# Short-Lived Hyperglycaemia Contributes to Continuous Glucose Monitoring Metrics Used for Therapeutic Decision-Making

**DOI:** 10.64898/2026.10.28.26365991

**Authors:** Guillaume Deside, Zain Syed, Laura Symul, Philippe Lysy

## Abstract

**Background:** Continuous glucose monitoring (CGM) metrics, including time in range (TIR) and median glucose, guide therapeutic decisions in type 1 diabetes (T1D) but do not distinguish sustained from transient hyperglycaemia. We investigated the prevalence of short-lived hyperglycaemia (SLH), i.e. brief isolated hyperglycaemic episodes, and its contribution to CGM metrics in T1D children and adolescents.

**Methods:** In this monocentric retrospective cohort study, T1D patients aged 6 months–20 years, enrolled in 2025–2026, with 5-minute CGM data, ≥70% coverage and median glucose 6·1–9·4 mmol/L within a 45-day window were included. SLH were hyperglycaemic episodes lasting 10–60 minutes surrounded by normoglycaemia, classified by onset as daytime or night-time. Their contribution to CGM metrics was estimated by replacing SLH with plausible trajectories; sensitivity analyses varied construction parameters.

**Findings:** Among 135 participants, 2,874 of 15,556 hyperglycaemic episodes (18·5%) met daytime SLH criteria (median 19·0 per participant). Removing SLH increased daytime TIR by 1·74 percentage points (95% CI 1·59–1·89) and reduced daytime median glucose by 0·18 mmol/L (95% CI 0·17–0·20). Whole-day effects were modest, but local impact was substantially greater: at maximal divergence, glucose profiles differed by 1·30 mmol/L (95% CI 1·21–1·38), around meals. Patients with near-identical conventional metrics could show markedly different SLH frequency.

**Interpretation:** SLH are common in this paediatric T1D cohort and alter reported CGM metrics: modest whole-day effects contrast with greater impact at specific times. Distinguishing transient from sustained hyperglycaemia provides clinically relevant information beyond aggregate metrics and may guide behavioural or educational interventions rather than insulin intensification.

**Funding:** Francqui Foundation and Helmsley Charitable Trust.

**Research in context:** *Evidence before this study:* We searched PubMed, Embase, and Web of Science from database inception to 31 March 2026, without language restriction, using the terms “short-lived hyperglycaemia” OR “transient hyperglycaemia” OR “brief hyperglycaemia” OR “isolated hyperglycaemia” OR “postprandial excursion” AND (“time in range” OR TIR OR “median glucose” OR “ambulatory glucose profile” OR CGM metric) AND (“type 1 diabetes” OR T1D) AND (paediatric OR children OR adolescents). We also reviewed reference lists of relevant reviews and ISPAD/ADA consensus statements on CGM interpretation. CGM metrics guide therapeutic decisions but do not distinguish sustained hyperglycaemia from brief, isolated excursions, and patients with similar metrics show heterogeneous glucose patterns. No study had quantified the contribution of such episodes to routine CGM metrics, and episode definitions were heterogeneous, without isolation criteria or a reproducible detection method.

*Added value of this study:* This is the first study to define short-lived hyperglycaemia (SLH) with explicit duration, isolation, and data-completeness criteria, and to quantify its contribution to CGM metrics using counterfactual glucose trajectories with these episodes removed. In this paediatric cohort, SLH accounted for about one-fifth of hyperglycaemic episodes; removing them increased daytime time in range by 1·74 percentage points, and median daily glucose profiles differed by up to 1·30 mmol/L around meals. SLH were more frequent in younger children, in those with shorter diabetes duration, and in those with better overall glycaemic control (lower GTAA1c, a score combining HbA1c and time in range). Patients with nearly identical conventional metrics differed markedly in SLH frequency, a temporal pattern invisible to summary metrics alone.

*Implications of all the available evidence:* Because recurrent SLH episodes can inflate the CGM metrics used for insulin titration, clinicians should check whether hyperglycaemia is brief and isolated before intensifying insulin, as behavioural or educational measures may then be more appropriate. Future studies should integrate automated SLH detection into CGM reports and test prospectively, with meal, insulin, and activity data, whether targeting SLH improves individualised management and reduces overtreatment.

## Introduction

Type 1 diabetes (T1D) is an autoimmune disease requiring lifelong insulin therapy [1], and optimal glycaemic control remains challenging, particularly in children and adolescents [2, 3]. Suboptimal control increases the risk of acute and long-term microvascular and macrovascular complications [4].

Glycated haemoglobin (HbA1c), the clinical gold standard, reflects average glucose over 2–3 months but does not capture short-term fluctuations or glycaemic variability [5–7]. Continuous glucose monitoring (CGM) provides standardised summary metrics, including mean and median glucose, time in range (TIR), and the coefficient of variation (CV), which are now central to therapeutic decision-making and international recommendations [8–10]. However, these metrics provide limited information on the temporal structure of hyperglycaemia: patients with similar TIR or median glucose may show sustained hyperglycaemia [11] or multiple brief postprandial excursions [12,13], patterns that likely reflect different mechanisms and require different therapeutic approaches.

In the present study, we focus on a specific pattern of transient hyperglycaemia that we term short-lived hyperglycaemia (SLH), defined as brief, isolated hyperglycaemic excursions that rapidly return to near-normoglycaemic levels. These episodes are common in otherwise well-controlled patients and are often attributed to delayed insulin administration, inaccurate carbohydrate estimation, or high-glycaemic-index snacks rather than persistent insulin deficiency. Nevertheless, when recurrent, these transient excursions may increase the glucose metrics displayed in routine CGM reports. Consequently, clinicians may interpret elevated mean or median glucose values as evidence of inadequate insulin dosing and intensify insulin therapy, whereas the underlying problem may instead require behavioural or educational interventions. Such therapeutic intensification may fail to address the cause of the excursions and potentially increase the risk of hypoglycaemia. Whether such episodes meaningfully influence routine CGM metrics, including robust measures such as median glucose, and how this varies across individuals remains unknown. We therefore developed algorithms to detect SLH episodes and construct counterfactual CGM trajectories in children and adolescents with T1D. Specifically, we aimed to (1) characterise the prevalence and temporal distribution of SLH episodes, (2) quantify their contribution to median glucose and other CGM metrics used for therapeutic decision-making, (3) assess inter-individual variability in this contribution, and (4) identify clinical characteristics associated with SLH frequency.

## Methods

### Study design and population

This monocentric, retrospective study characterised SLH episodes in children and adolescents with type 1 diabetes. The study was conducted in the Paediatric Diabetes Centre of the Cliniques universitaires Saint-Luc (CUSL, Brussels, Belgium). All procedures were conducted in accordance with the Declaration of Helsinki. The study protocol was approved by the Comité d’éthique hospitalo-facultaire des Cliniques universitaires Saint-Luc (protocol 2022/02FEV/043). Patient enrolment and the analysis phase extended from March 2025 to May 2026.

Patients were eligible if they were aged between 6 months and 20 years and diagnosed with diabetes according to the American Diabetes Association (ADA) criteria (American Diabetes Association Professional Practice 2023) defined by at least one of the following: haemoglobin A1C ≥ 6·5%, fasting plasma glucose (FPG) ≥ 7 mmol/L, two-hour plasma glucose during a 75-g oral glucose tolerance test (OGTT) ≥ 11·1 mmol/L or random plasma glucose ≥ 11·1 mmol/L accompanied by classic hyperglycaemic symptoms or hyperglycaemic crises. To be included in the analysis, patients were required to be under CGM (using Freestyle Libre 2 or 3®, Abbott) and have CGM data collected at a 5-minute resolution for at least three consecutive months. Patients also needed at least one 45-day window with ≥70% data coverage, based on prior recommendations [14]. When multiple 45-day windows met this criterion, we selected the window with the highest coverage for analysis. Patients with median glucose outside 6·1–9·4 mmol/L in the analysis window were excluded, as this likely reflects unmanaged diabetes.

In addition to the CGM data, clinical variables were extracted from the consultation visit occurring within the three-month window of CGM data. Clinical variables extracted from the medical record included age, sex, diabetes duration, HbA1c, insulin regimen and dose, BMI z-score [15], insulin-dose-adjusted HbA1c (IDAA1c), glycaemic target-adjusted HbA1c (GTAA1c), and history of DKA or severe hypoglycaemia.

### Definition and detection of SLH episode

We defined SLH episodes as short, isolated peaks > 10·0 mmol/L that are physiologically distinct from sustained hyperglycaemia. Specifically, an episode was labelled SLH if it met three criteria:

1. <u>Duration of hyperglycaemia</u>: the duration of the hyperglycaemia episode (> 10 mmol/L) had to be of at least 10 minutes and at most 60 minutes.
2. <u>Near-normal glycaemia (< 10·0 mmol/L) in</u> the 60 min before and after, with at least one reading < 7·8 mmol/L: the CGM readings had to be smaller than 10 mmol/L for the 60 minutes preceding and following the hyperglycaemia episodes, and at least one reading < 7·8 mmol/L in that window.
3. <u>Data completeness</u> during and around the hyperglycaemia episode: the data coverage during the episode and within 60 minutes of the episode had to be at least 75%.

Episodes passing all three criteria were classified as SLH. Those that passed the first criterion but not the last two were classified as non-isolated SLH. All remaining episodes were classified as long-lived hyperglycaemia (LLH). Non-isolated SLH and LLH episodes are collectively referred to as non-SLH episodes. Among SLH episodes, we differentiated daytime *vs.* night-time episodes based on their onset time. SLH episodes starting after 06:00 and before 22:00 were classified as daytime SLH and as night-time SLH otherwise. This distinction reflects our assumption that nocturnal hyperglycaemia is most likely due to basal insulin inadequacy rather than meal-related excursions.

### Counterfactual signal construction

Consistent with the study’s underlying hypothesis that SLH episodes are primarily driven by suboptimal eating behaviours (*e.g.,* snacking on high glycaemic index food) or suboptimal timing of insulin doses, and that SLH episodes may then contribute to inflating CGM summary metrics used for clinical decision on insulin dosing, we sought to quantify that contribution by constructing *counterfactual* CGM time series. These counterfactual time series correspond to the glucose trajectories that we assumed would have been observed if the underlying cause had not occurred.

In practice, we redrew each episode as a straight line from the lowest glucose just before it to the lowest just after it, asking what the CGM trace would look like without that excursion. We computed these counterfactual trajectories by removing daytime SLH episodes and interpolating between anchor points surrounding the episodes. The anchor points are the minimal CGM values measured within a context window (δ, in minutes) surrounding the SLH onset and offset. The main analyses were performed using a 30-minute window, while sensitivity analyses used windows of δ = 0, 15, 45, 60, 75, and 90 minutes. Night-time SLH were not removed and remained unaffected by the procedure. The full mathematical description of the counterfactual construction is provided in Supplementary methods S1 (Counterfactual signal construction; equations S1–S4).

### Contribution of SLH episodes to summary metrics

The effect of SLH episode removal on CGM summary metrics (median and mean glucose, TIR 3·9–10·0, CV, SD, TAR >10 and >13·9, and TBR <3·9) was quantified as the difference between the metrics computed on the observed *vs.* counterfactual glycaemia. Changes were summarised across participants as the mean and 95% confidence interval (CI) around the mean.

We computed two versions of these summary metrics. The “24-h” metrics were calculated over the full glucose signal, using all 24 hours of each day, as routinely reported in clinical practice. The “daytime” metrics were calculated over the daytime period (06:00–22:00) of each day (using the same daytime definition as for classifying daytime *vs.* night-time SLH).

In addition to these aggregate metrics, we quantified the local contribution of SLH episodes to the daily glucose profile. Specifically, for each participant, we computed the median observed and median counterfactual glucose profiles at each time of day over the 45 days of the analysis window, and their difference Δm(t). We then reported t_max_, the time of day at which |Δm(t)| was maximal, and the corresponding maximum absolute difference |Δm(t_max_)| (in mmol/L). Formal definitions are given in Supplementary methods S2 (Local contribution of SLH episodes to the daily glucose profile; equations S5–S9).

### Association between SLH episode numbers and baseline characteristics

We quantified associations between the number of SLH episodes and three groups of variables. Baseline CGM metrics included mean and median glucose, SD, CV, TIR 3·9–10·0 mmol/L, and TAR >10·0 mmol/L. Clinical phenotype variables comprised continuous variables (age at study entry, diabetes duration, BMI z-score, and total daily insulin dose) and categorical variables (sex, insulin regimen [basal-bolus therapy, insulin pump therapy, or two daily injections], presence of at least one comorbidity, family history of diabetes, reported physical activity, diabetic ketoacidosis (DKA) at diagnosis, DKA after diagnosis, and history of severe hypoglycaemia). Glycaemic biomarkers included HbA1c, IDAA1c, and GTAA1c.

Associations between the number of SLH episodes and these variables were assessed using Spearman rank correlation, given the non-normal, over-dispersed distribution of SLH counts. For categorical phenotype variables, SLH counts were compared across categories using the Mann–Whitney U test for binary variables and the Kruskal–Wallis test for variables with more than two categories. We performed a sensitivity analysis using *mutual information*, a measure of any kind of association, not only monotonic ones. Pairwise correlations among all predictors were also computed to highlight collinearity within and across domains. Software and packages used for all analyses are listed in the Supplementary material.

## Results

### Cohort characteristics

Out of 366 patients registered in the CUSL paediatric diabetes convention, 245 had at least 3 consecutive months of CGM data, and 135 met all eligibility criteria (**Figure 1**; Figure S1). Cohort characteristics and baseline CGM metrics are in Table 1.

**Figure 1:**
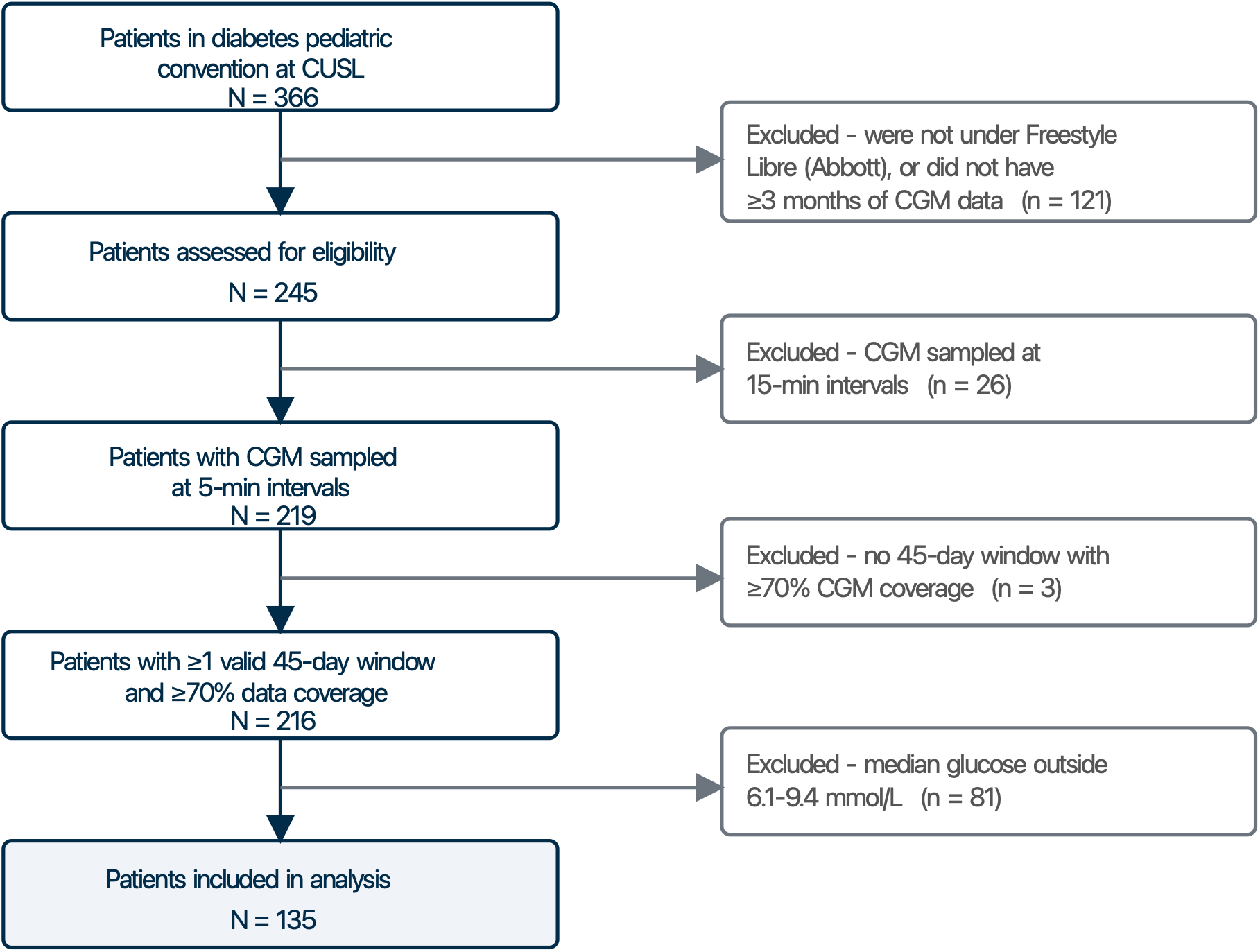
Study flow chart. Of the 366 patients registered in the CUSL paediatric diabetes convention, 245 had at least three consecutive months of continuous glucose monitoring data and 135 met all eligibility and inclusion criteria.

**Table 1:**
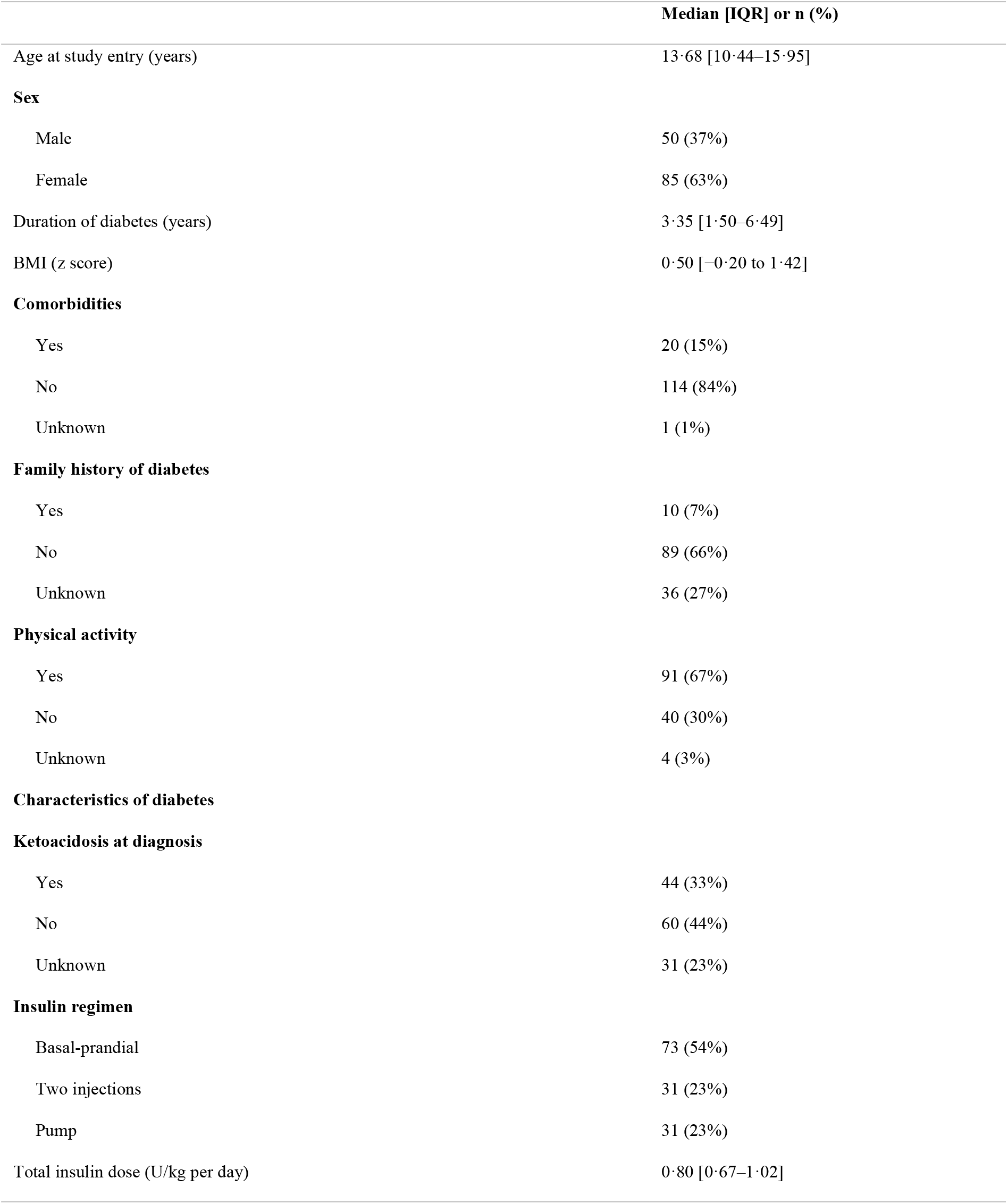
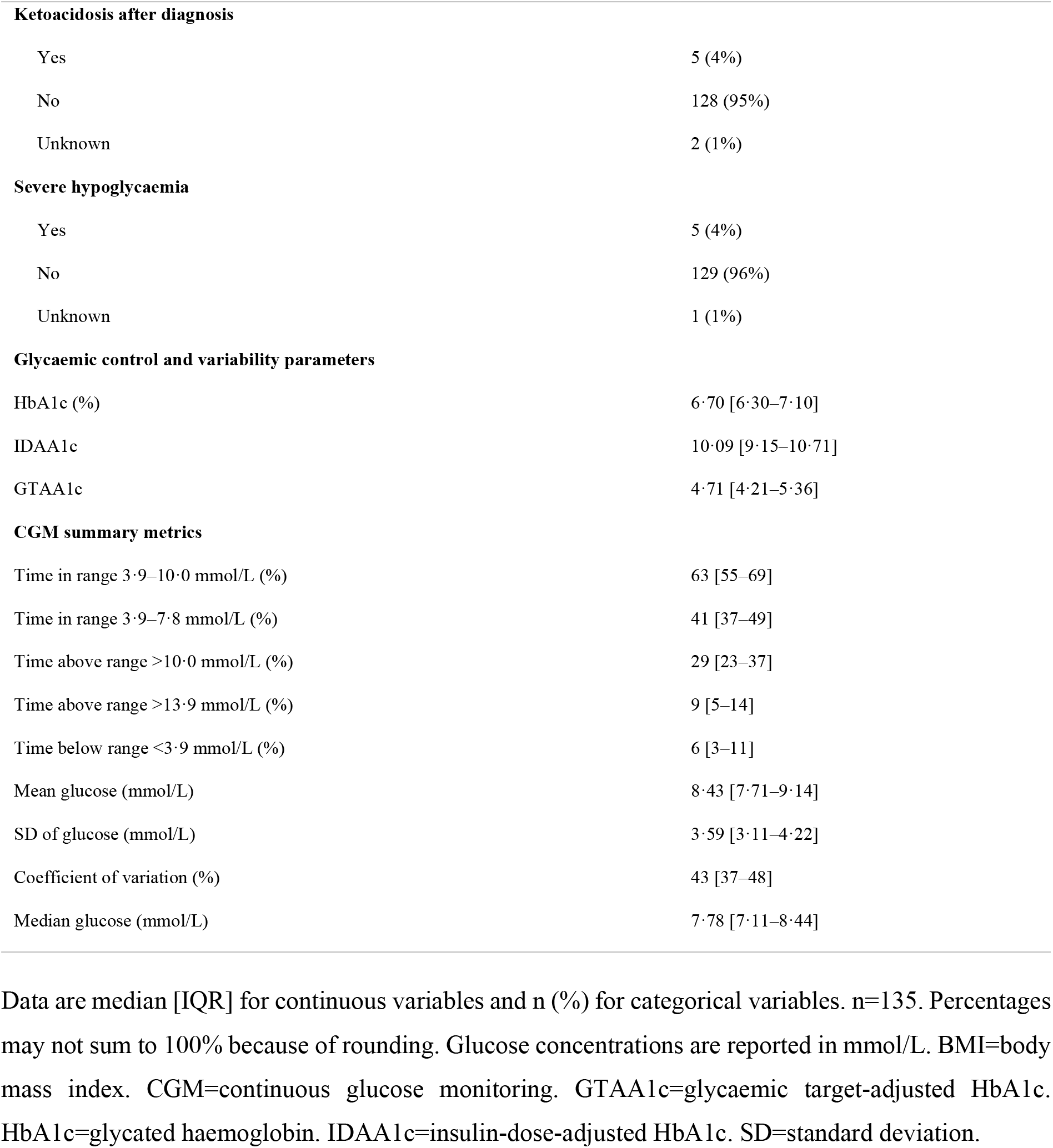
Characteristics of the study cohort.

|  | Median [IQR] or n (%) |
|---|---|
| Age at study entry (years) | 13·68 [10·44–15·95] |
| **Sex** |  |
| Male | 50 (37%) |
| Female | 85 (63%) |
| Duration of diabetes (years) | 3·35 [1·50–6·49] |
| BMI (z score) | 0·50 [−0·20 to 1·42] |
| **Comorbidities** |  |
| Yes | 20 (15%) |
| No | 114 (84%) |
| Unknown | 1 (1%) |
| **Family history of diabetes** |  |
| Yes | 10 (7%) |
| No | 89 (66%) |
| Unknown | 36 (27%) |
| **Physical activity** |  |
| Yes | 91 (67%) |
| No | 40 (30%) |
| Unknown | 4 (3%) |
| **Characteristics of diabetes** |  |
| **Ketoacidosis at diagnosis** |  |
| Yes | 44 (33%) |
| No | 60 (44%) |
| Unknown | 31 (23%) |
| **Insulin regimen** |  |
| Basal-prandial | 73 (54%) |
| Two injections | 31 (23%) |
| Pump | 31 (23%) |
| Total insulin dose (U/kg per day) | 0·80 [0·67–1·02] |
| **Ketoacidosis after diagnosis** |  |
| Yes | 5 (4%) |
| No | 128 (95%) |
| Unknown | 2 (1%) |
| **Severe hypoglycaemia** |  |
| Yes | 5 (4%) |
| No | 129 (96%) |
| Unknown | 1 (1%) |
| **Glycaemic control and variability parameters** |  |
| HbA1c (%) | 6·70 [6·30–7·10] |
| IDAA1c | 10·09 [9·15–10·71] |
| GTAA1c | 4·71 [4·21–5·36] |
| **CGM summary metrics** |  |
| Time in range 3·9–10·0 mmol/L (%) | 63 [55–69] |
| Time in range 3·9–7·8 mmol/L (%) | 41 [37–49] |
| Time above range >10·0 mmol/L (%) | 29 [23–37] |
| Time above range >13·9 mmol/L (%) | 9 [5–14] |
| Time below range <3·9 mmol/L (%) | 6 [3–11] |
| Mean glucose (mmol/L) | 8·43 [7·71–9·14] |
| SD of glucose (mmol/L) | 3·59 [3·11–4·22] |
| Coefficient of variation (%) | 43 [37–48] |
| Median glucose (mmol/L) | 7·78 [7·11–8·44] |

Data are median [IQR] for continuous variables and n (%) for categorical variables. n=135. Percentages may not sum to 100% because of rounding. Glucose concentrations are reported in mmol/L. BMI=body mass index. CGM=continuous glucose monitoring. GTAA1c=glycaemic target-adjusted HbA1c. HbA1c=glycated haemoglobin. IDAA1c=insulin-dose-adjusted HbA1c. SD=standard deviation.

### Short-lived hyperglycaemia episodes are frequent

Across the 45-day analysis window of the 135 patients, 15,556 hyperglycaemic episodes (>10·0 mmol/L) were detected. Of these, 5,480 (35·2%) met the duration criterion and 3,237 were classified as SLH, of which 363 were night-time SLH and 2,874 (18·5%) were classified as daytime SLH (**Figure 2A**). Examples of each classification outcome are shown in Figure S2, and a representative participant trace is shown in Figures S3 and S4.

**Figure 2:**
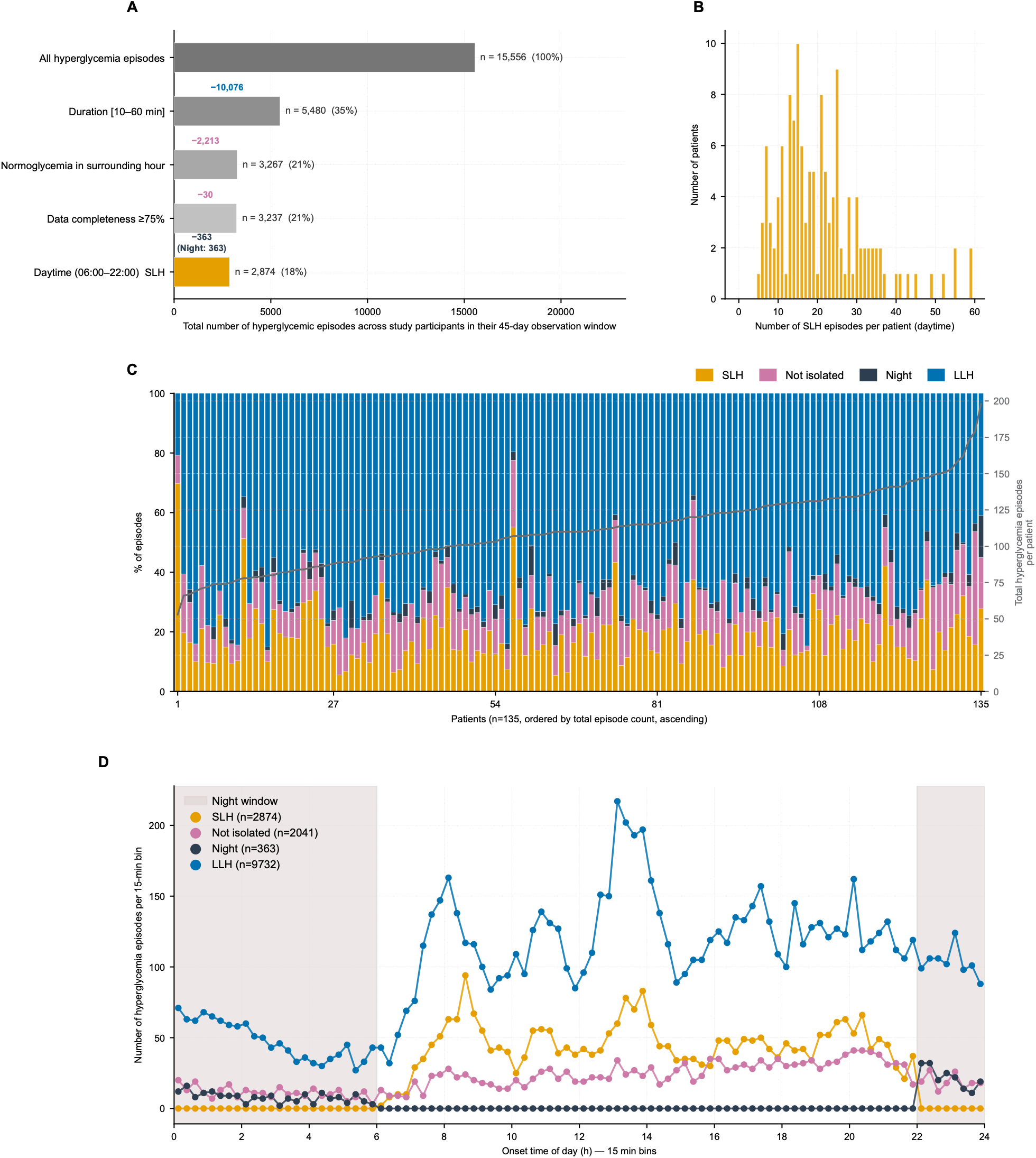
Classification of hyperglycaemic episodes. (A) Waterfall of episodes remaining after each classification criterion (duration, context, completeness, daytime), ending in the short-lived hyperglycaemia (SLH) count. (B) Episodes excluded at each criterion. (C) Number of daytime SLH episodes per patient. (D) Episode onset time of day, by category.

There was a median of 19·0 daytime SLH episodes per patient (IQR 13·0–25·5; range 5–59; Figure 2B, C), versus a median of 2·0 for night-time SLH (IQR 1·0–4·0; range 0–28). The proportion of daytime SLH among all hyperglycaemic episodes varied greatly between patients, from 5·3% to 69·8% (median 17·0%; Figure 2C). Overall, both LLH and SLH episodes were more frequent at expected meal/snack times (**Figure 2D**). However, the time distribution of SLH and LLH episodes varied among participants, with some patients experiencing SLH but no LLH episodes at specific times of day (**Figure S5**).

### SLH frequency is variable across patients

To illustrate this heterogeneity, two patients with similar CGM summary metrics but large differences in SLH numbers were selected: patient A174 with 59 SLH episodes and patient A224 with 19 (**Figure 3**). A174 also had more non-isolated SLH and LLH episodes combined (83 *vs.* 58; 142 vs. 77 episodes in total). Despite this, their CGM summary metrics were nearly identical (**Figure 3G**): TAR >10·0 mmol/L was 17·2% *vs.* 17·1% and median glucose 7·1 *vs*. 6·9 mmol/L.

**Figure 3:**
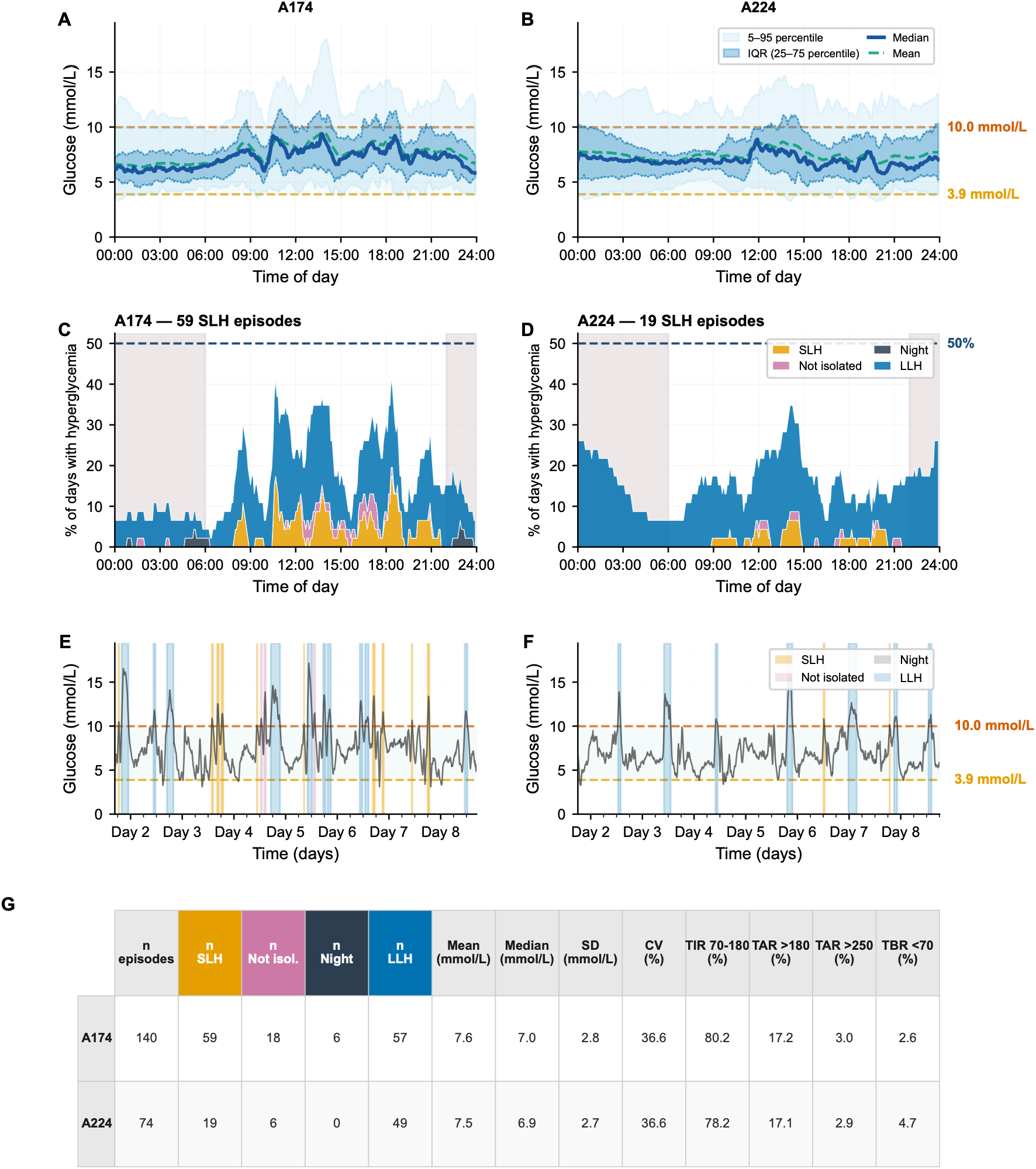
Glucose profiles and short-lived hyperglycaemia (SLH) patterns in two representative patients with similar median glucose (patient A174, 59 SLH episodes; patient A224, 19). (A–B) Ambulatory glucose profiles (median, IQR, and 5th–95th percentiles). (C–D) SLH episodes per time-of-day bin across the 45-day window. (E–F) Representative 7-day glucose traces; hyperglycaemia threshold (10·0 mmol/L) in red. (G) Standard continuous glucose monitoring metrics and SLH and long-lived hyperglycaemia (LLH) episode counts for both patients. IQR=interquartile range.

### In silico *intervention quantifies* the contribution of SLH episodes to CGM summary metrics

In these two patients, our in-silico intervention (counterfactual glucose trajectories without SLH; **Figure 4A,B**; Figures S6, S7) yielded different results (**Figure 4C–F**). For patient A174, the intervention led to an estimated daytime TIR gain of 4·68 percentage points and a daytime median glucose reduction of −0·33 mmol/L (**Figure 4E, F**). In contrast, A224 had a more modest daytime TIR improvement of 1·50 percentage points and no change in median glucose (**Figure 4E, F**). Improvements in TIR were robust to different choices of counterfactual construction parameters while improvement in median glucose levels increased with the strength of SLH removal (**Figure 4E, F**).

**Figure 4:**
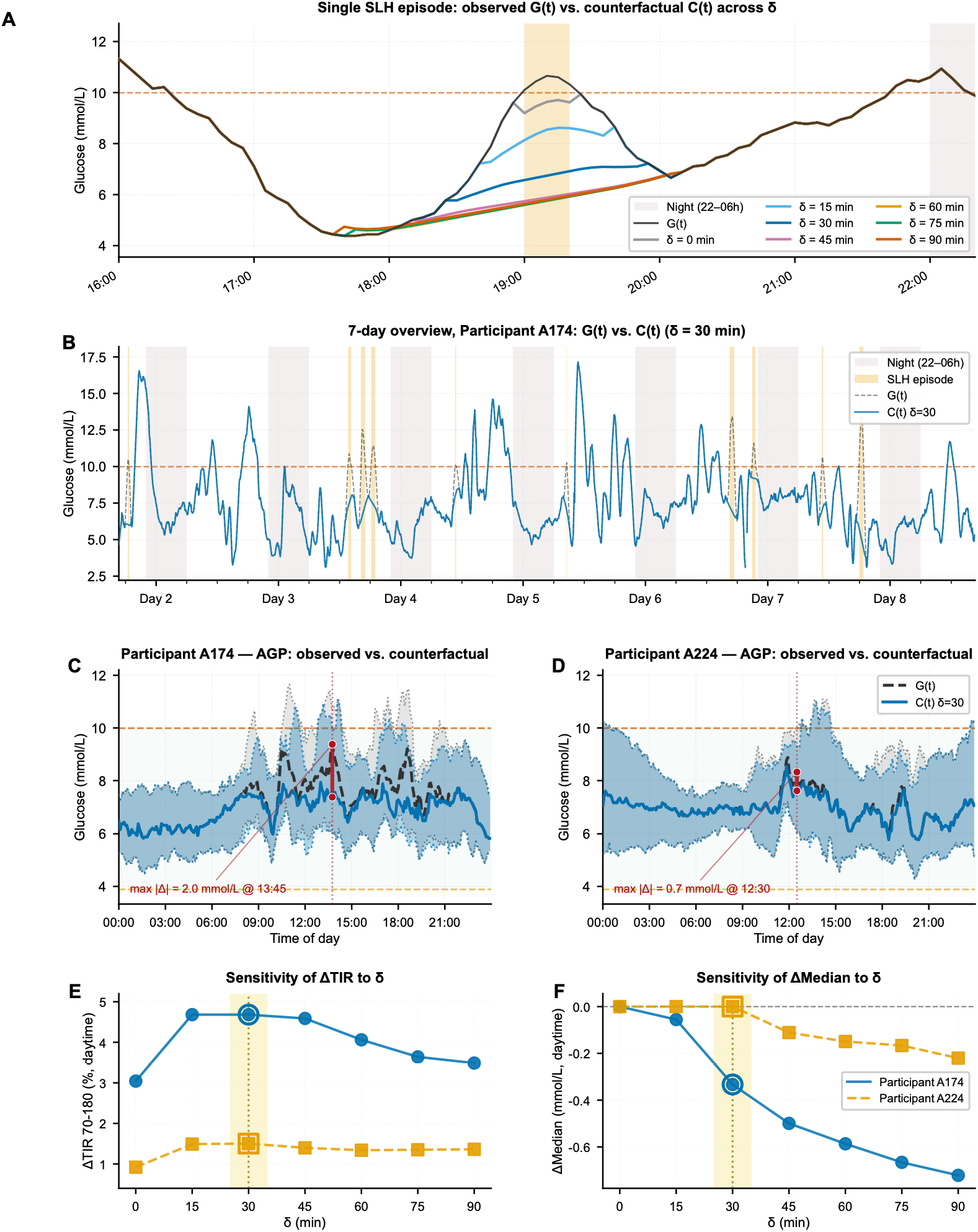
Counterfactual glucose trajectories for the two patients shown in Figure 3 (A174 and A224; δ=30 min unless stated). (A) Counterfactual construction for one SLH episode (patient A224) for each context window δ tested (0–90 min). (B) Observed and counterfactual glucose over 7 days (patient A174). (C, D) Ambulatory glucose profiles (median and IQR) before and after SLH removal in patients A174 (C) and A224 (D). (E, F) Change in daytime time in range (E) and median glucose (F) after SLH removal by context window width; shaded band, main analysis. IQR=interquartile range. SLH=short-lived hyperglycaemia.

Eliminating their SLH episodes also had a drastically different impact on their median daily profile. While the time at which the median profile most diverged from their counterfactual was similar (13:45 for A174 and 12:30 for A224), the maximum difference between the two median glucose profiles reached 2·00 mmol/L for A174 and only 0·72 mmol/L for A224 (**Figure 4C, D**).

### Contribution of SLH episodes in the whole cohort

Across the cohort of 135 patients, counterfactual removal of SLH episodes produced a mean improvement in daytime TIR of 1·74 percentage points (95% CI 1·59–1·89; Figure 5A) and a mean reduction in daytime median glucose of 0·18 mmol/L (95% CI 0·17–0·20; Figure 5B). Effects on all other summary metrics, including 24-h values, are provided in Table 2.

**Figure 5:**
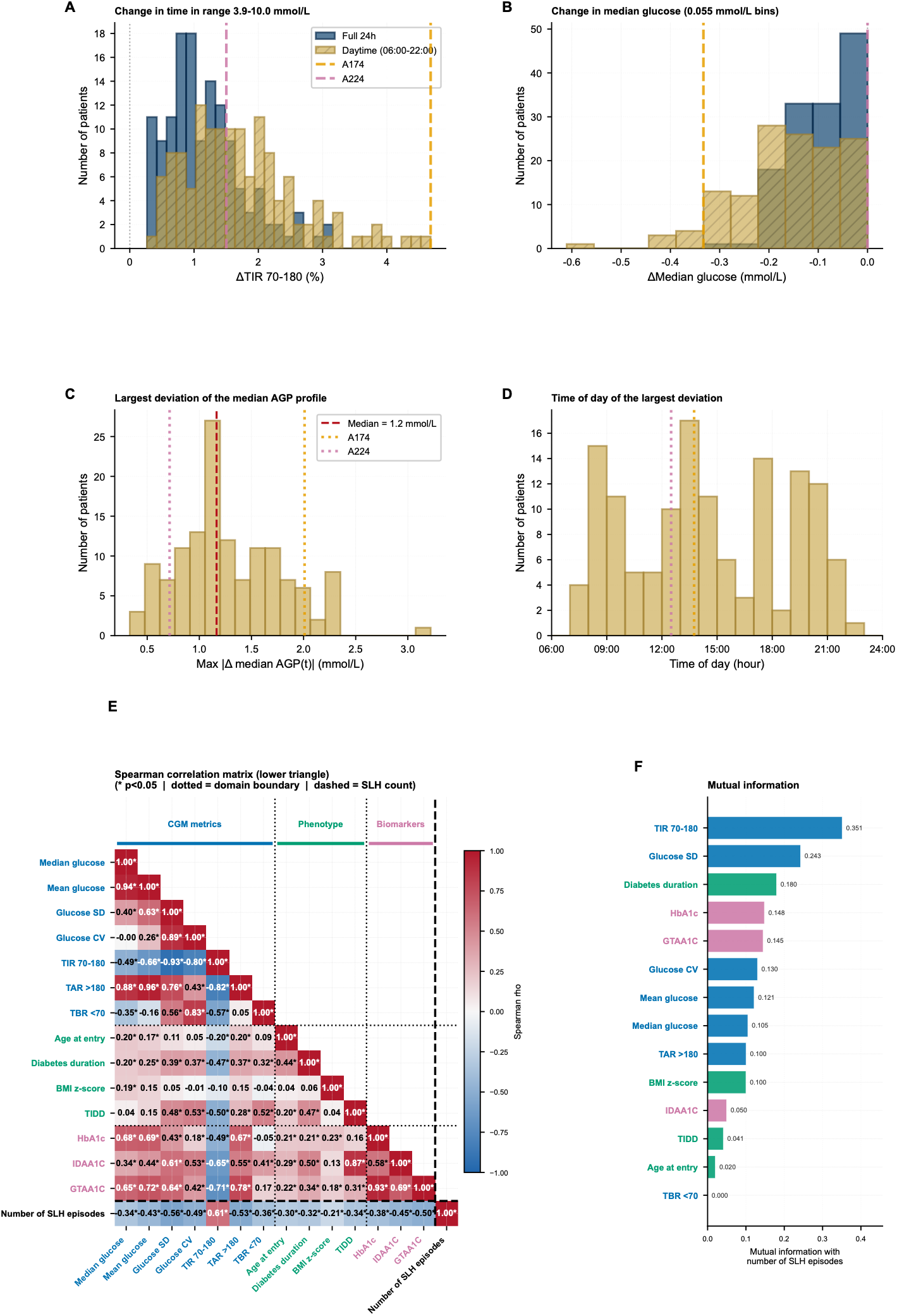
Cohort-level contribution of short-lived hyperglycaemia (SLH) episodes to continuous glucose monitoring metrics and baseline correlates of SLH frequency (n = 135, δ = 30 min). (A) Change in daytime and 24-h time in range after SLH removal. (B) Change in daytime and overall median glucose. (C) Maximum difference between the observed and counterfactual median glucose profiles. (D) Time of day at which the maximum difference occurred. (E) Spearman correlation and mutual information between baseline glucose metrics and SLH number. SLH=short-lived hyperglycaemia.

**Table 2:** Change in continuous glucose monitoring summary metrics after in silico removal of short-lived hyperglycaemia (SLH) episodes.

|  | **Analysis period** | **δ = 30** |
|---|---|---|
| Time above range >10·0 mmol/L (%) | 24-h | −1·18 [−1·28, −1·07] |
| Time above range >10·0 mmol/L (%) | Daytime | −1·74 [−1·90, −1·59] |
| Time above range >13·9 mmol/L (%) | 24-h | 0·00 [−0·01, 0·00] |
| Time above range >13·9 mmol/L (%) | Daytime | −0·01 [−0·01, 0·00] |
| Time below range <3·9 mmol/L (%) | 24-h | 0·00 [0·00, 0·00] |
| Time below range <3·9 mmol/L (%) | Daytime | 0·00 [0·00, 0·01] |
| Time in range 3·9–10·0 mmol/L (%) | 24-h | 1·17 [1·07, 1·28] |
| Time in range 3·9–10·0 mmol/L (%) | Daytime | 1·74 [1·59, 1·89] |
| Mean glucose (mmol/L) | 24-h | −0·07 [−0·07, −0·06] |
| Mean glucose (mmol/L) | Daytime | −0·10 [−0·11, −0·09] |
| Median glucose (mmol/L) | 24-h | −0·12 [−0·13, −0·10] |
| Median glucose (mmol/L) | Daytime | −0·18 [−0·20, −0·17] |
| Coefficient of variation (%) | 24-h | 0·17 [0·12, 0·23] |
| Coefficient of variation (%) | Daytime | 0·31 [0·23, 0·39] |
| SD of glucose (mmol/L) | 24-h | −0·01 [−0·02, −0·01] |
| SD of glucose (mmol/L) | Daytime | −0·01 [−0·02, −0·01] |

Data are mean [95% CI] change from the observed to the counterfactual signal for 135 patients, at a context window of δ = 30 min (results for all context windows are shown in Table S1). Negative values indicate a reduction after removal of short-lived hyperglycaemia episodes. Time in range, time above range, time below range, and coefficient of variation are expressed as percentage points. Glucose concentrations are reported in mmol/L. CGM=continuous glucose monitoring. SD=standard deviation. CI=confidence interval.

As expected by construction, the improvement in 24-h TIR was strongly correlated with the number of SLH episodes (ρ = 0·98). In contrast, the improvement in daytime median glucose level was less correlated with the number of SLH episodes (ρ = −0·35).

Median glucose profiles after SLH removal diverged most from their original profiles by a mean |Δm(t_max_)| of 1·30 mmol/L (95% CI 1·21–1·38; Table 3), most often after typical mealtimes (modes at 09:00, 13:00, 17:00, 20:00; **Figure 5C, D).**

**Table 3:** Difference between observed and counterfactual median glucose profiles.

|  | **δ = 30** |
|---|---|
| Maximum absolute difference between daily median glucose profiles, \|Δm(t<sub>max</sub>)\| (mmol/L) | 1·30 [1·21, 1·38] |
| Time of maximum difference, t<sub>max</sub> (h) | 14:30 [13:46, 15:14] |

Data are mean [95% CI] for \|Δm(tmax)\| and median [IQR] for tmax, for 135 patients, at a context window of δ = 30 min (results for all context windows are shown in Table S2). Δm(t) is the difference, at time of day t, between the observed and counterfactual median glucose profiles; \|Δm(t<sub>max</sub>)\| is the maximum absolute difference and t<sub>max</sub> the time of day at which it occurs, expressed as 24-h clock time (hh:mm). Glucose concentrations are reported in mmol/L. CI=confidence interval.

### Sensitivity analyses suggest results are robust to parameter choices

We evaluated the effects of SLH removal across seven context window widths (δ = 0–90 min in 15-min steps; Tables S1–S2; Figure S8). TIR improvement was largest at δ = 15–30 min and declined modestly at wider windows, in parallel with a small increase in TBR, but remained positive across all windows. Improvements in mean and median glucose increased with δ, but the increments diminished at the widest windows.

### Baseline characteristics associated with SLH number

Because the number of SLH episodes was strongly associated with the effect of their removal on CGM summary metrics (Figure S9), we assessed whether some patients were more likely than others to experience many SLH episodes.

The CGM metrics were significantly associated with SLH number (*all p*<0·05; **Figure 5E**). TIR 3·9–10 mmol/L was the strongest correlate (ρ = +0·61). Glucose variability metrics (SD, TAR >10·0 mmol/L, CV) and, more weakly, central-tendency metrics (mean, median glucose) correlated negatively.

Glycaemic variability indexes were also associated with SLH frequency. GTAA1c was the strongest non-CGM correlate (ρ = −0·49, *p<0·001*), higher values corresponding to fewer SLH episodes.

SLH number decreased with diabetes duration (ρ = −0·32, p<0·001) and age (ρ = −0·30, p<0·001); GTAA1c, in turn, increased with diabetes duration (ρ = +0·34, *p<0·001*; Figure S10). Among categorical phenotype variables, only insulin regimen was associated with SLH number (Kruskal–Wallis p=0·008; Figure S10).

## Discussion

In this retrospective study of 135 children and adolescents with type 1 diabetes, we identified a distinct pattern of glycaemic excursions that we termed SLH. Using a predefined three-criterion classification algorithm and a counterfactual signal reconstruction framework, we quantified their contribution to commonly used CGM-derived metrics. First, SLH episodes were frequent, accounting for approximately one-fifth of all hyperglycaemic episodes. Second, removal of SLH episodes improved CGM summary metrics, particularly TIR and median glucose, as well as median daily profiles, indicating that these excursions, although brief, contribute substantially to overall glycaemic profiles. Third, patients with similar conventional CGM metrics exhibited very different glucose temporal dynamics, suggesting these patients may have different underlying hyperglycaemic phenotypes or drivers.

The increasing adoption of CGM has transformed diabetes management by providing metrics that extend beyond HbA1c, including TIR, TAR, and measures of glycaemic variability. These metrics are now central to therapeutic decision-making and are recommended by both ISPAD and international consensus guidelines for paediatric diabetes care [5,8,16]. However, standard CGM metrics aggregate glucose values across time and generally do not distinguish between different temporal patterns of hyperglycaemia. As a result, prolonged hyperglycaemia caused by inadequate insulin therapy and brief postprandial excursions both contribute to summary statistics despite potentially reflecting different underlying mechanisms and requiring different therapeutic responses.

Our findings suggest that recurrent SLH episodes represent a clinically relevant source of heterogeneity not adequately captured by conventional CGM interpretation. Two patients may present comparable median glucose, TAR, or TIR values while exhibiting fundamentally different glycaemic dynamics. Our representative patient comparison illustrates this: despite nearly identical baseline CGM metrics, patients differed markedly in SLH frequency and consequently in the estimated benefit obtained after removal of these episodes. This observation supports the concept that CGM-derived summary measures alone may provide an incomplete description of glycaemic behaviour and highlights the potential value of incorporating temporal pattern analysis into routine CGM interpretation.

By construction, SLH events were isolated excursions, predominantly with a daytime onset, that returned rapidly towards normoglycaemia and were not preceded or followed by sustained hyperglycaemia. Such patterns are consistent with transient postprandial glucose rises, potentially associated with high-glycaemic-index snacks, inaccuracies in carbohydrate counting, delayed insulin administration, or other behavioural factors. Unlike prolonged hyperglycaemia, which often reflects persistent insulin insufficiency, these excursions may not necessarily indicate a need for insulin intensification. Yet, in routine CGM reports, frequent SLH episodes may raise mean or median glucose and prompt clinicians to intensify insulin therapy when behavioural or educational interventions would be more appropriate. This distinction is clinically important because therapeutic escalation based solely on elevated TAR or reduced TIR could increase the risk of hypoglycaemia if the underlying problem is behavioural rather than pharmacological [10,14].

Methodologically, we proposed a reproducible classification framework that isolates brief excursions from the broader spectrum of hyperglycaemic events. The inclusion of contextual criteria, particularly the requirement for glucose values <7·8 mmol/L before or after the episode, was intended to identify excursions demonstrating physiological recovery rather than progression towards sustained dysglycaemia. This approach extends existing CGM analyses, which generally rely on threshold-based metrics without considering the temporal structure of individual glucose excursions [7,10,14]. The algorithm was intentionally conservative, retaining only episodes meeting all criteria, which increases specificity.

The counterfactual framework represents another novel aspect of the study. Rather than merely counting episodes, we estimated their contribution to CGM metrics by reconstructing glucose trajectories without SLH episodes. Although this approach cannot establish causality, it provides a clinically intuitive estimate of the extent to which transient excursions influence commonly reported CGM outcomes. Sensitivity analyses showed that results were robust to interpolation parameters. Wider windows increased the estimated gain in glucose levels but reduced the gain in TIR, which may reflect that, even when the time above 10 mmol/L is short, many SLH episodes are part of longer excursions from lower baseline levels.

Interestingly, patients with higher SLH frequency also had better baseline glycaemic control, including higher TIR and lower measures of glycaemic variability. This apparent paradox should be interpreted in the context of our highly selected study population, which was restricted to children with relatively well-controlled diabetes and excluded individuals with marked hypo- or hyperglycaemia. Within such a cohort, prolonged hyperglycaemic episodes were intentionally underrepresented, making isolated postprandial excursions account for a larger proportion of overall hyperglycaemic exposure. Consequently, individuals spending most of their time within the target range may accumulate a greater relative number of SLH episodes despite otherwise favourable glycaemic control. This interpretation is consistent with previous observations that identical TIR values may arise from markedly different glycaemic patterns and reinforces the importance of evaluating glucose dynamics beyond aggregate metrics [7,12,13].

Besides CGM-derived baseline metrics, GTAA1c was the strongest clinical correlate of SLH frequency, with higher values associated with fewer episodes. SLH frequency also decreased with diabetes duration. These findings suggest that isolated SLH episodes are more common in children with shorter disease duration and relatively preserved glycaemic control. This pattern may reflect greater susceptibility to meal-related glycaemic excursions and may identify a phenotype more amenable to behavioural interventions; however, meal data were not available to test this hypothesis.

Several limitations should be acknowledged. First, the study was retrospective and conducted at a single tertiary paediatric diabetes centre, which may limit generalisability. Second, the interpretation of SLH episodes as behaviour-related excursions remains hypothetical because information regarding meal timing, carbohydrate intake, physical activity, and insulin administration was not available. Third, the counterfactual reconstruction relies on interpolation assumptions and therefore represents an analytical estimate rather than a direct observation of glucose trajectories. Alternative modelling approaches could, however, yield different estimates. Finally, we did not assess associations with patient-reported outcomes, treatment modifications, or long-term clinical endpoints.

Future research should focus on validating the SLH interpretation and effects in independent cohorts and exploring its applicability in adults with T1D and individuals with type 2 diabetes. Integrating meal, insulin, and activity data may help identify the behavioural and physiological determinants of SLH episodes and determine whether targeted educational interventions reduce their occurrence. Additionally, the development of automated CGM reporting tools that distinguish transient from sustained hyperglycaemic patterns may improve clinical interpretation and support more individualised treatment decisions.

In conclusion, SLH episodes are common in children and adolescents with type 1 diabetes and contribute measurably to standard CGM-derived metrics. Conventional glycaemic metrics do not distinguish between transient and sustained forms of hyperglycaemia, potentially obscuring clinically relevant differences in glucose dynamics. Our classification and counterfactual framework offers a novel way to quantify the influence of transient excursions and may complement existing CGM interpretation strategies. Recognition of SLH patterns could help clinicians better differentiate behavioural from therapeutic contributors to dysglycaemia and thereby support more individualised diabetes management.

## Data Availability

Individual participant data (de-identified CGM recordings and clinical data) will not be made publicly available, as data sharing is not covered by the ethics approval and the consent obtained for this study. Data may be considered upon reasonable request to the corresponding author, subject to ethics committee approval and a data use agreement. The analysis code is publicly available at https://github.com/gdeside/counterfactual-cgm-slh.

## Contributions

PL conceived the study and provided clinical oversight. PL and ZS curated the clinical and CGM data. GD and ZS designed the short-lived hyperglycaemia classification algorithm and the counterfactual framework. GD did the statistical analysis. GD wrote the first draft of the manuscript (writing – original draft). LS supervised the study (supervision). LS, ZS, and PL critically reviewed and edited the manuscript (writing – review and editing).

## Data sharing

Individual participant data, including de-identified CGM recordings and clinical data, will be made available from the corresponding author upon reasonable request.

The analysis code (SLH detection and counterfactual construction) is available at https://github.com/gdeside/counterfactual-cgm-slh and archived on Zenodo (https://doi.org/10.5281/zenodo.23615326).

## Acknowledgements

We thank all the children, adolescents, and young adults with type 1 diabetes and their families whose data made this study possible. We are grateful to the physicians, diabetes nurses, dietitians, psychologists, and administrative staff of the paediatric diabetes convention at the Cliniques universitaires Saint-Luc for their clinical care of participants and their help with data collection.

GD’s salary is covered by a Seed Grant awarded to LS by the Francqui Foundation.

Artificial intelligence tools were used to assist with language editing, reformulation of selected passages, and formatting of tables. All scientific content, analyses, interpretations, and conclusions remain the sole responsibility of the authors.

