## Supplementary material for "Short-Lived Hyperglycaemia Contributes to Continuous Glucose Monitoring Metrics Used for Therapeutic Decision-Making"

#### Supplementary methods

##### S1. Counterfactual signal construction

Let $G(t)$ denote the observed CGM signal of a participant over the 45-day analysis window and let $\mathcal{L}$ denote the set of daytime SLH episodes detected in that window. Each episode $\ell \in \mathcal{L}$ is characterised by its onset $O_\ell$ and offset $E_\ell$, and $\delta \geq 0$ denotes the length of the context windows preceding the onset and following the offset ($\delta = 30$ min in the main analysis; $\delta \in \{0, 15, 45, 60, 75, 90\}$ min in sensitivity analyses). For each episode, a left anchor $L_a = (t_a, g_a)$ and a right anchor $L_b = (t_b, g_b)$ are defined as the time and value of the minimum glucose reading in the left context window $[O_\ell - \delta, O_\ell]$ and in the right context window $[E_\ell, E_\ell + \delta]$, respectively:

$$t_a = \underset{t \in [O_\ell - \delta, O_\ell]}{\arg\min}\ G(t), \quad g_a = G(t_a) \tag{S1}$$

$$t_b = \underset{t \in [E_\ell, E_\ell + \delta]}{\arg\min}\ G(t), \quad g_b = G(t_b) \tag{S2}$$

The SLH episode is then replaced by the straight line joining the two anchors, while the signal is left unchanged outside the interval $[t_a, t_b]$:

$$\tilde{C}(t) = \begin{cases} g_a + (g_b - g_a)\frac{t - t_a}{t_b - t_a}, & \text{if } t_a \leq t \leq t_b \\ G(t), & \text{otherwise} \end{cases} \tag{S3}$$

Finally, within each interpolated interval $[t_a, t_b]$, the signal is smoothed with a centred rolling-mean filter of kernel width $k = 60$ min:

$$C(t) = \frac{1}{n_k(t)} \sum_{s \in W_k(t)} \tilde{C}(s), \quad W_k(t) = \left[t - \frac{k}{2}, t + \frac{k}{2}\right] \tag{S4}$$

where $n_k(t)$ is the number of CGM readings in $W_k(t)$. Equations (S1)–(S4) are applied episode by episode to every $\ell \in \mathcal{L}$. Night-time SLH episodes are not processed, so that $C(t) = G(t)$ outside the daytime SLH intervals.

#### S2. Local contribution of SLH episodes to the daily glucose profile

To quantify the local effect of SLH removal on the daily glucose profile, the observed and counterfactual signals were organised by day and time of day. Let $G_d(t)$ and $C_d(t)$ denote the observed and counterfactual glucose values on day $d \in \{1, \ldots, D\}$ ($D = 45$) at time of day $t$. The median observed and counterfactual daily profiles are computed at each time of day as:

$$m_G(t) = \underset{d=1,\ldots,D}{\operatorname{median}} G_d(t) \tag{S5}$$

$$m_C(t) = \underset{d=1,\ldots,D}{\operatorname{median}} C_d(t) \tag{S6}$$

and the local contribution of SLH episodes at time of day $t$ is defined as their difference:

$$\Delta m(t) = m_G(t) - m_C(t) \tag{S7}$$

For each participant, we then identified the time of day at which this difference is largest in absolute value, together with the corresponding signed difference:

$$t_{\max} = \arg\max_{t} |\Delta m(t)| \tag{S8}$$

$$\Delta m(t_{\max}) = m_G(t_{\max}) - m_C(t_{\max}) \tag{S9}$$

The maximal difference $\Delta m(t_{\max})$ is expressed in mmol/L and $t_{\max}$ as 24-h clock time (hh:mm).

#### Software and packages

Analyses were performed using Python 3.11.5 and the following packages: pandas 2.1.0 for data manipulation, numpy 1.24.3 for numerical operations, scipy 1.11.2 for statistical testing, scikit-learn 1.8.0 for mutual information estimation, matplotlib 3.7.2, and seaborn 0.12.2 for visualisations. Analysis code and data processing pipelines are available at https://github.com/gdeside/counterfactual-cgm-slh and archived on Zenodo (https://doi.org/10.5281/zenodo.23615326).

### Figure S1

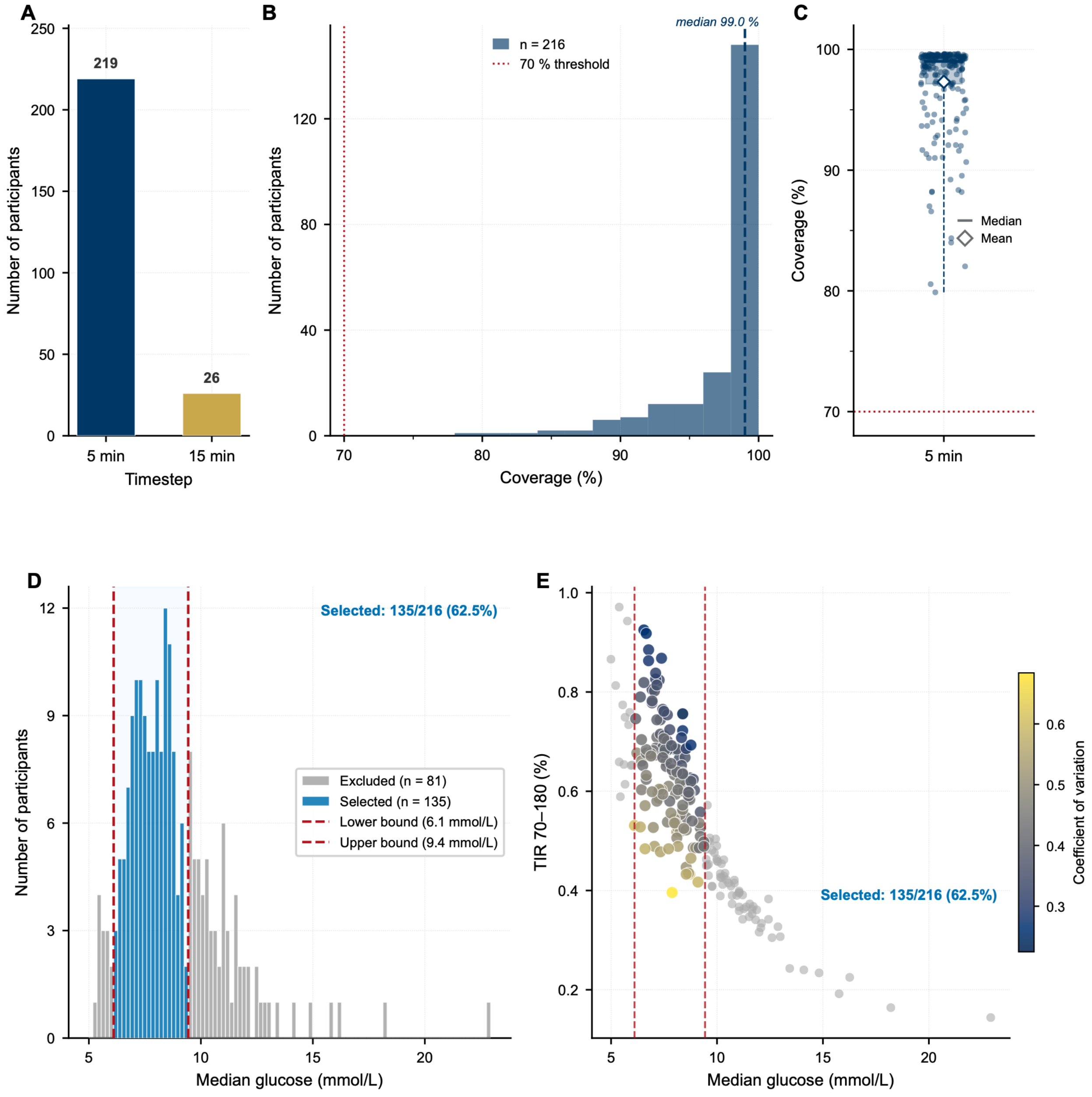


**Figure S1:** Data quality and cohort selection. (A) Dominant continuous glucose monitoring (CGM) sampling intervals across all 245 participants. (B) Data coverage for participants with 5-min sampling and coverage above 70% (n = 216). (C) Individual coverage: median (line), mean (diamond), IQR (box), and range (whiskers). (D) Median glucose distribution (n = 216). Blue bars and shaded region: selected participants (median glucose 6·1–9·4 mmol/L, dashed lines); grey bars: excluded. (E) Median glucose against time in range 3·9–10·0 mmol/L, coloured by coefficient of variation; dashed lines mark the selection boundaries.

### Figure S2

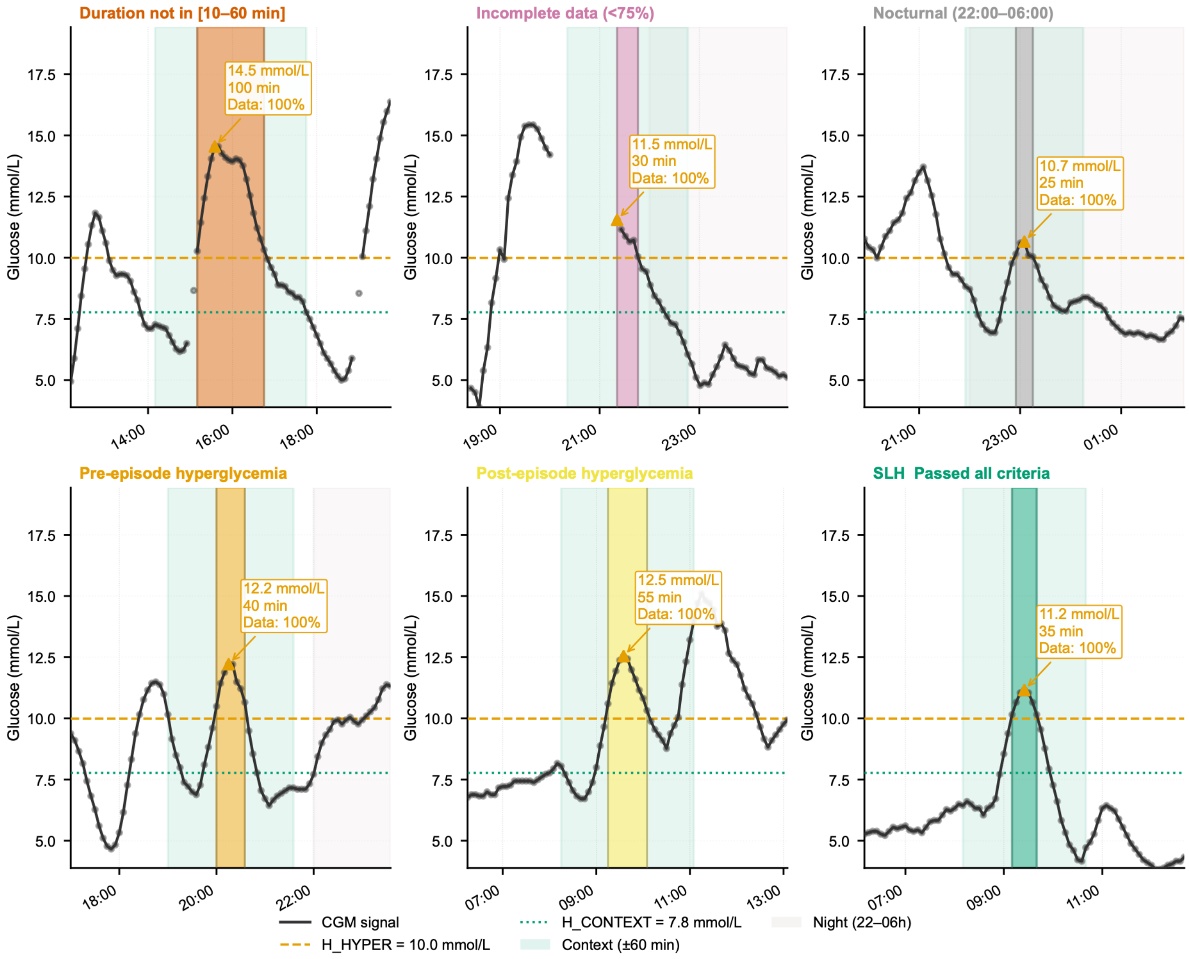


**Figure S2:** Six individual episodes from participant A159 illustrating each classification outcome in the sequential filtering pipeline. Each panel displays a 3–6 h window centred on the episode, with the hyperglycaemic excursion highlighted in colour corresponding to its classification status. Black line represents the CGM trace G(t); orange dashed line marks the hyperglycaemia threshold (10·0 mmol/L); blue dotted line indicates the context threshold (7·8 mmol/L). Light teal shading denotes the contextual windows (±60 min before and after episode boundaries); grey shading indicates night-time periods (22:00–06:00).

### Figure S3

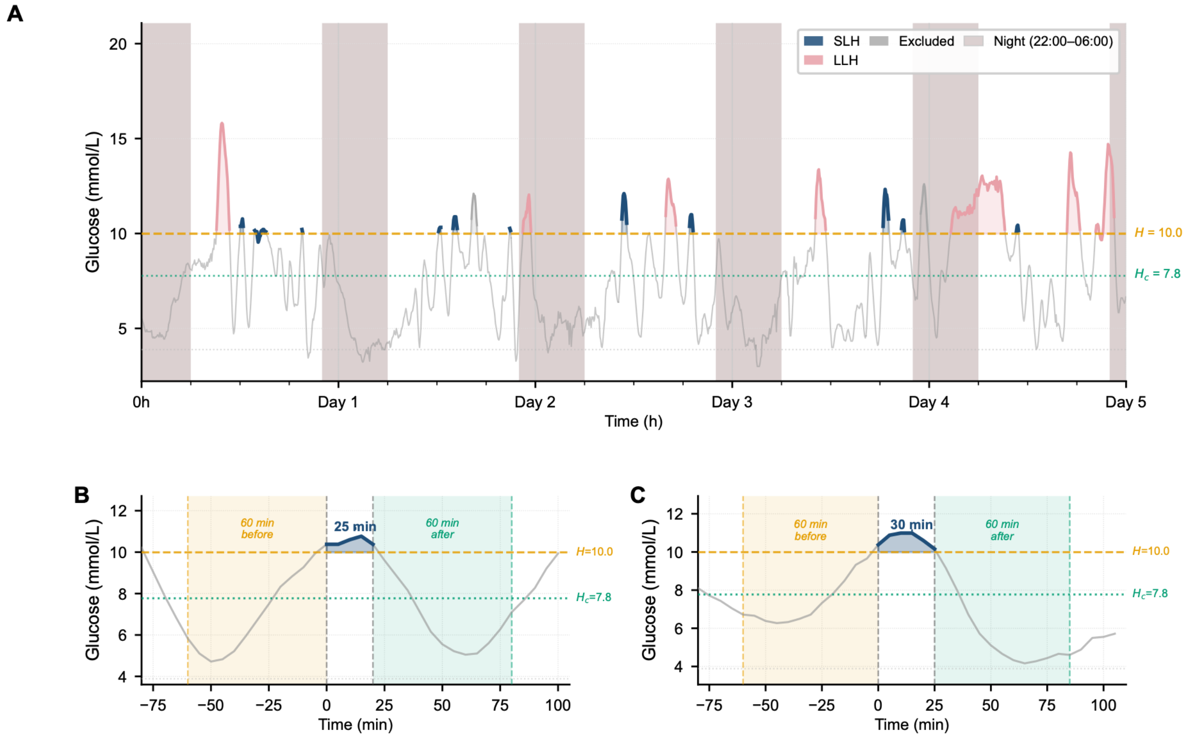


**Figure S3:** Representative example from participant A174. (A) Five-day continuous glucose monitoring trace showing all detected hyperglycaemic episodes (>10·0 mmol/L) colour-coded by final classification. Navy shading indicates confirmed SLH episodes; pink shading indicates long-lived hyperglycaemia (LLH, n=57); grey shading indicates episodes excluded as non-isolated (n=26). Together, LLH and non-isolated episodes account for the 83 non-SLH episodes for this participant. Grey vertical bands mark night-time periods (22:00–06:00). Horizontal dashed line: hyperglycaemia threshold (10·0 mmol/L, orange). Horizontal dotted line: context threshold (7·8 mmol/L, teal). (B–C) Detailed views of two representative SLH episodes.

### Figure S4

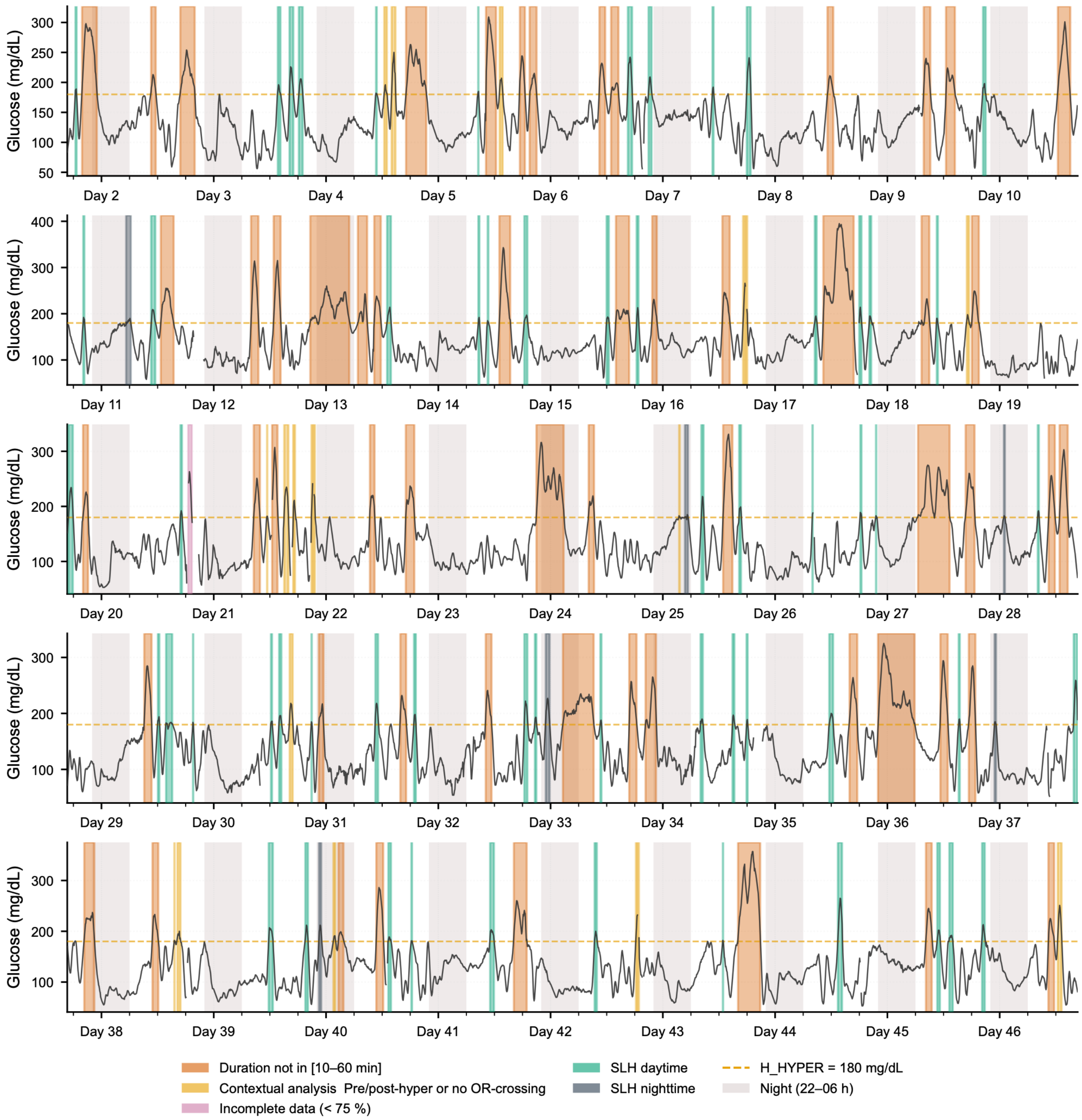


**Figure S4:** Complete glucose trace for participant A174 with all detected hyperglycaemic episodes colour-coded according to their classification status.

### Figure S5

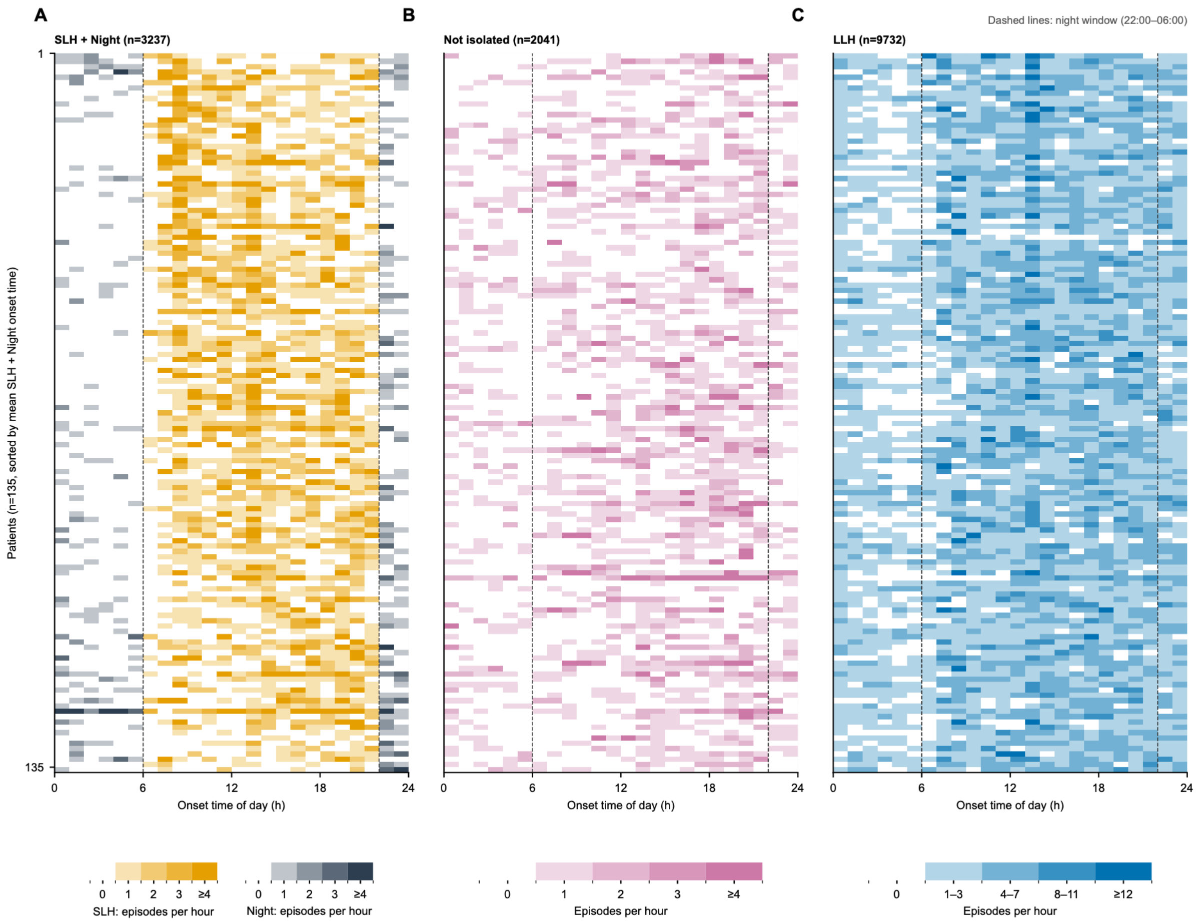


**Figure S5:** Time of day at which hyperglycaemic episodes start, for each participant and each episode category. Each panel shows all participants (rows, n = 135), with the same participant on the same row in every panel. Columns are hours of the day, and the colour intensity is the number of episodes that started in that hour (0 to ≥4). Panels show (A) daytime and night-time short-lived hyperglycaemia (SLH) episodes, (B) non-isolated SLH episodes, and (C) long-lived hyperglycaemia (LLH). Participants are ordered by the mean onset time of their SLH episodes, followed by participants without SLH. Dashed lines mark the night window (22:00–06:00). The time distribution of SLH and LLH varies between participants, and some participants have SLH but no LLH at specific times of day.

#### Figure S6

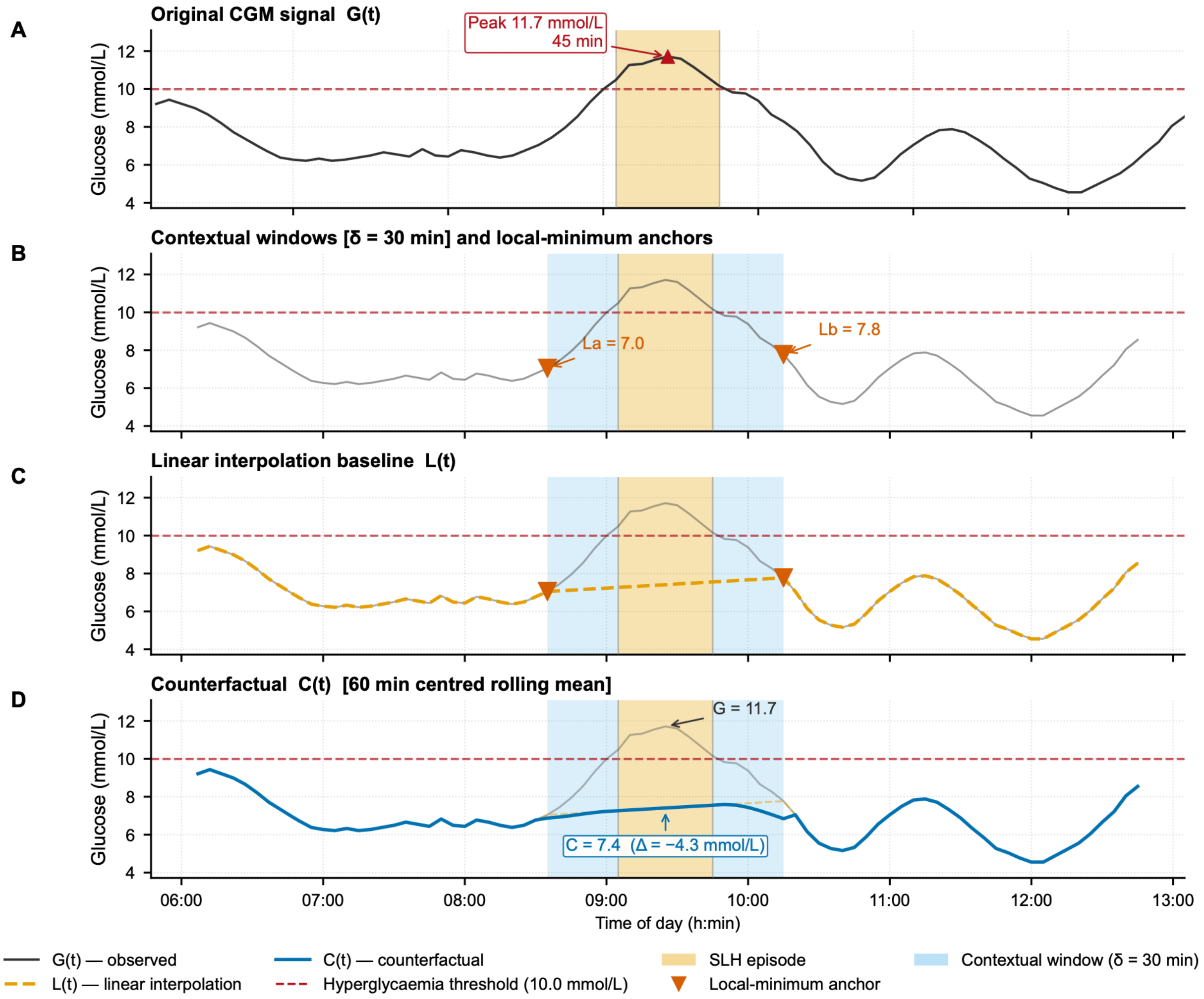


**Figure S6:** Counterfactual signal construction for one daytime short-lived hyperglycaemia (SLH) episode (participant A159; onset 09:05, duration 45 min, peak 11·7 mmol/L; ±3 h of context shown). (A) Observed signal G(t), with peak (triangle) and episode above the hyperglycaemia threshold (shaded; 10·0 mmol/L, dashed line). (B) Context windows (δ=30 min, paler bands) and their minima La and Lb (inverted triangles). (C) Linear interpolation $\tilde{C}(t)$ between La and Lb. (D) Counterfactual C(t) after 60 min centred rolling-mean smoothing, lying 4·3 mmol/L below the observed peak.

### Figure S7

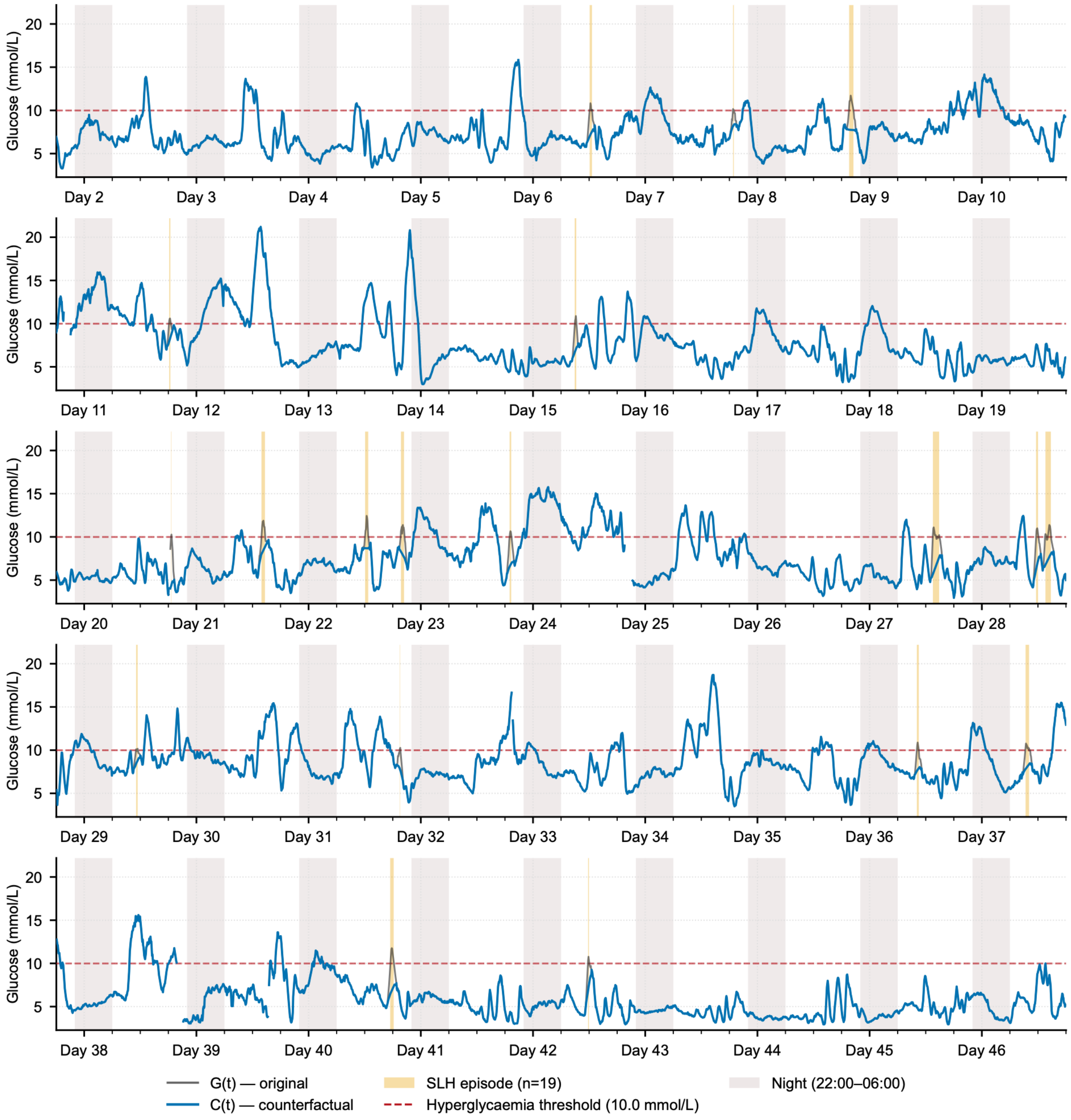


**Figure S7:** Complete 45-day observation window for participant A224 showing original CGM trace G(t) and counterfactual signal C(t) constructed using recommended parameters (δ = 30 min context window, k = 60 min rolling mean).

### Table S1

**Table S1: Change in continuous glucose monitoring summary metrics after in silico removal of short-lived hyperglycaemia (SLH) episodes**

| | **Analysis period** | **δ = 0** | **δ = 15** | **δ = 30** | **δ = 45** | **δ = 60** | **δ = 75** | **δ = 90** |
|---|---|---|---|---|---|---|---|---|
| Time above range >10·0 mmol/L (%) | 24-h | −0·74 [−0·80, −0·67] | −1·18 [−1·28, −1·07] | −1·18 [−1·28, −1·07] | −1·18 [−1·28, −1·07] | −1·18 [−1·28, −1·07] | −1·18 [−1·28, −1·07] | −1·18 [−1·28, −1·07] |
| Time above range >10·0 mmol/L (%) | Daytime | −1·09 [−1·19, −1·00] | −1·74 [−1·90, −1·59] | −1·74 [−1·90, −1·59] | −1·74 [−1·90, −1·59] | −1·74 [−1·90, −1·59] | −1·74 [−1·90, −1·59] | −1·74 [−1·90, −1·59] |
| Time above range >13·9 mmol/L (%) | 24-h | 0·00 [−0·01, 0·00] | 0·00 [−0·01, 0·00] | 0·00 [−0·01, 0·00] | 0·00 [−0·01, 0·00] | 0·00 [−0·01, 0·00] | 0·00 [−0·01, 0·00] | 0·00 [−0·01, 0·00] |
| Time above range >13·9 mmol/L (%) | Daytime | −0·01 [−0·01, 0·00] | −0·01 [−0·01, 0·00] | −0·01 [−0·01, 0·00] | −0·01 [−0·01, 0·00] | −0·01 [−0·01, 0·00] | −0·01 [−0·01, 0·00] | −0·01 [−0·01, 0·00] |
| Time below range <3·9 mmol/L (%) | 24-h | 0·00 [0·00, 0·00] | 0·00 [0·00, 0·00] | 0·00 [0·00, 0·00] | 0·05 [0·03, 0·06] | 0·13 [0·09, 0·16] | 0·24 [0·18, 0·29] | 0·36 [0·28, 0·44] |
| Time below range <3·9 mmol/L (%) | Daytime | 0·00 [0·00, 0·00] | 0·00 [0·00, 0·00] | 0·00 [0·00, 0·01] | 0·07 [0·05, 0·09] | 0·19 [0·14, 0·23] | 0·34 [0·26, 0·42] | 0·52 [0·40, 0·63] |
| Time in range 3·9–10·0 mmol/L (%) | 24-h | 0·74 [0·67, 0·80] | 1·18 [1·07, 1·28] | 1·17 [1·07, 1·28] | 1·13 [1·03, 1·23] | 1·05 [0·95, 1·15] | 0·94 [0·84, 1·04] | 0·82 [0·71, 0·92] |
| Time in range 3·9–10·0 mmol/L (%) | Daytime | 1·09 [1·00, 1·19] | 1·74 [1·59, 1·90] | 1·74 [1·59, 1·89] | 1·67 [1·52, 1·83] | 1·56 [1·41, 1·70] | 1·40 [1·25, 1·54] | 1·23 [1·07, 1·38] |
| Mean glucose (mmol/L) | 24-h | −0·01 [−0·01, −0·01] | −0·04 [−0·04, −0·03] | −0·07 [−0·07, −0·06] | −0·09 [−0·10, −0·08] | −0·11 [−0·12, −0·10] | −0·12 [−0·13, −0·11] | −0·14 [−0·15, −0·12] |
| Mean glucose (mmol/L) | Daytime | −0·02 [−0·02, −0·02] | −0·06 [−0·06, −0·05] | −0·10 [−0·11, −0·09] | −0·13 [−0·15, −0·12] | −0·16 [−0·17, −0·14] | −0·18 [−0·20, −0·17] | −0·20 [−0·22, −0·18] |
| Median glucose (mmol/L) | 24-h | 0·00 [0·00, 0·00] | −0·04 [−0·05, −0·03] | −0·12 [−0·13, −0·10] | −0·17 [−0·18, −0·15] | −0·20 [−0·21, −0·18] | −0·22 [−0·24, −0·21] | −0·24 [−0·26, −0·22] |
| Median glucose (mmol/L) | Daytime | 0·00 [−0·01, 0·00] | −0·07 [−0·09, −0·06] | −0·18 [−0·20, −0·17] | −0·25 [−0·28, −0·23] | −0·30 [−0·32, −0·27] | −0·33 [−0·36, −0·30] | −0·36 [−0·39, −0·33] |
| Coefficient of variation (%) | 24-h | −0·06 [−0·08, −0·04] | −0·02 [−0·06, 0·02] | 0·17 [0·12, 0·23] | 0·39 [0·32, 0·45] | 0·58 [0·50, 0·66] | 0·76 [0·67, 0·85] | 0·92 [0·82, 1·02] |
| Coefficient of variation (%) | Daytime | −0·07 [−0·10, −0·04] | 0·01 [−0·05, 0·06] | 0·31 [0·23, 0·39] | 0·63 [0·53, 0·73] | 0·92 [0·80, 1·04] | 1·18 [1·05, 1·32] | 1·42 [1·27, 1·56] |

| | | | | | | | | |
|---|---|---|---|---|---|---|---|---|
| SD of glucose (mmol/L) | 24-h | −0·01 [−0·01, −0·01] | −0·02 [−0·02, −0·01] | −0·01 [−0·02, −0·01] | 0·00 [−0·01, 0·00] | 0·00 [0·00, 0·01] | 0·01 [0·01, 0·02] | 0·02 [0·01, 0·03] |
| SD of glucose (mmol/L) | Daytime | −0·01 [−0·02, −0·01] | −0·02 [−0·03, −0·02] | −0·01 [−0·02, −0·01] | 0·00 [−0·01, 0·01] | 0·01 [0·01, 0·02] | 0·03 [0·02, 0·03] | 0·04 [0·03, 0·05] |

Data are mean [95% CI] change from the observed to the counterfactual signal for 135 patients, across context windows (δ, in minutes). Negative values indicate a reduction after removal of short-lived hyperglycaemia episodes. Time in range, time above range, time below range, and coefficient of variation are expressed as percentage points. Glucose concentrations are reported in mmol/L. CGM=continuous glucose monitoring. SD=standard deviation. CI=confidence interval.

### Table S2

##### Table S2: Difference between observed and counterfactual median glucose profiles

| | **δ = 0** | **δ = 15** | **δ = 30** | **δ = 45** | **δ = 60** | **δ = 75** | **δ = 90** |
|---|---|---|---|---|---|---|---|
| Maximum absolute difference between daily median glucose profiles, $\|\Delta m(t_{max})\|$ (mmol/L) | 0·50 [0·43, 0·56] | 0·98 [0·90, 1·06] | 1·30 [1·21, 1·38] | 1·46 [1·37, 1·55] | 1·55 [1·46, 1·65] | 1·62 [1·52, 1·71] | 1·65 [1·56, 1·75] |
| Time of maximum difference, $t_{max}$ (h) | 12:45 [11:35, 13:55] | 14:35 [13:49, 15:20] | 14:30 [13:46, 15:14] | 14:10 [13:23, 14:55] | 14:10 [13:25, 14:56] | 14:16 [13:25, 15:01] | 14:31 [13:46, 15:16] |

Data are mean [95% CI] for |Δm(tmax)| and median [IQR] for tmax, for 135 patients, across context windows (δ, in minutes). Δm(t) is the difference, at time of day t, between the observed and counterfactual median glucose profiles; $|\Delta m(t_{max})|$ is the maximum absolute difference and $t_{max}$ the time of day at which it occurs, expressed as 24-h clock time (hh:mm). Glucose concentrations are reported in mmol/L. CI=confidence interval.

### Figure S8

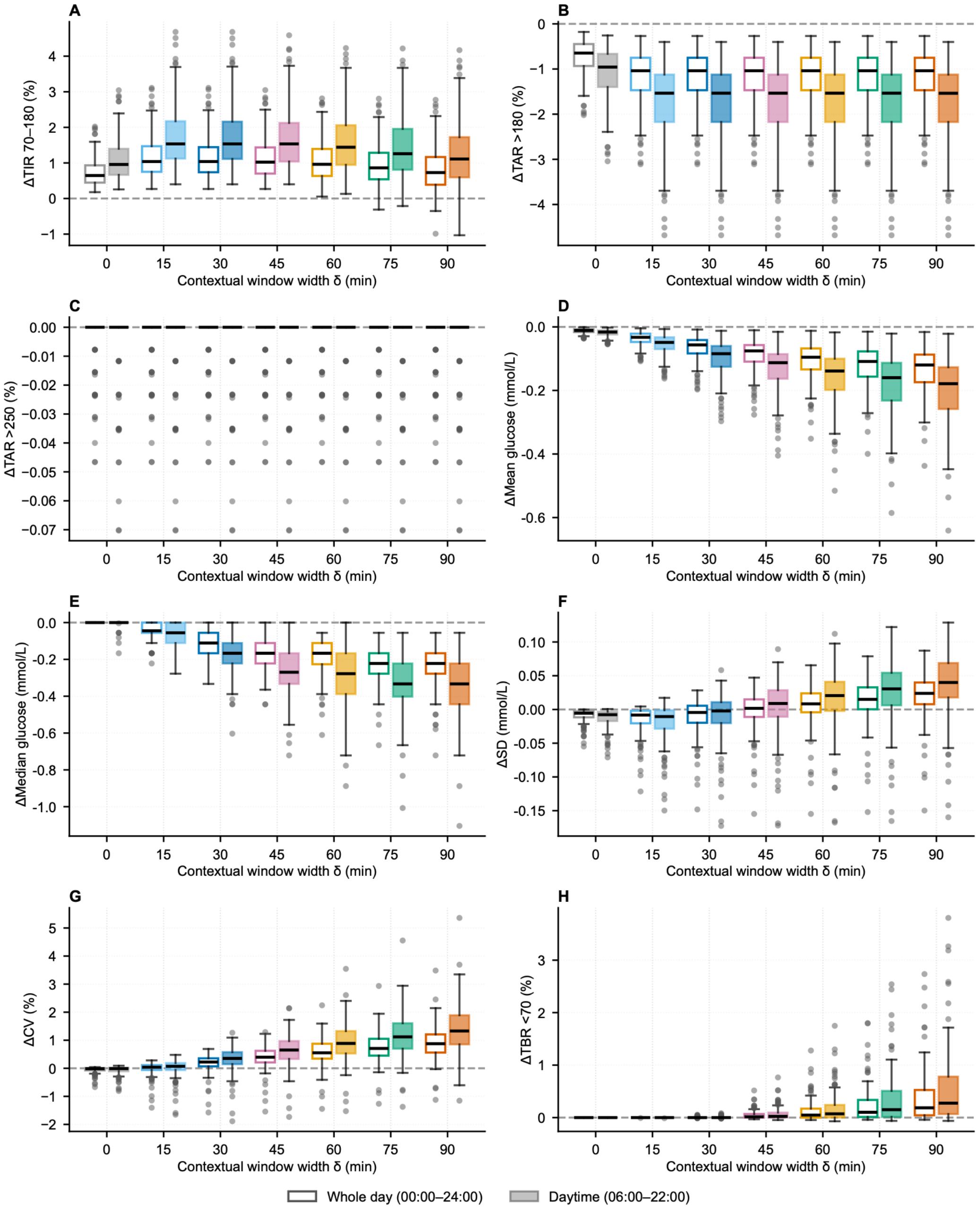


**Figure S8:** Sensitivity of cohort CGM summary metrics to the context window width δ. Per-participant change (n=135) after in silico removal of short-lived hyperglycaemia (SLH) episodes, for each δ tested (0–90 min, 15-min steps): (A) ΔTIR 3·9–10·0 mmol/L; (B) ΔTAR >10·0 mmol/L; (C) ΔTAR >13·9 mmol/L; (D) ΔMean glucose; (E) ΔMedian glucose; (F) ΔSD; (G) ΔCV; (H) ΔTBR <3·9 mmol/L (percentage points for TIR, TAR, TBR and CV; mmol/L otherwise). Open boxes: whole day; solid boxes: daytime (06:00–22:00); colour encodes δ. Counterfactual signals were built as described in Supplementary methods S1. Boxes: median and IQR; whiskers: 1·5 × IQR; dashed line: no change.

### Figure S9

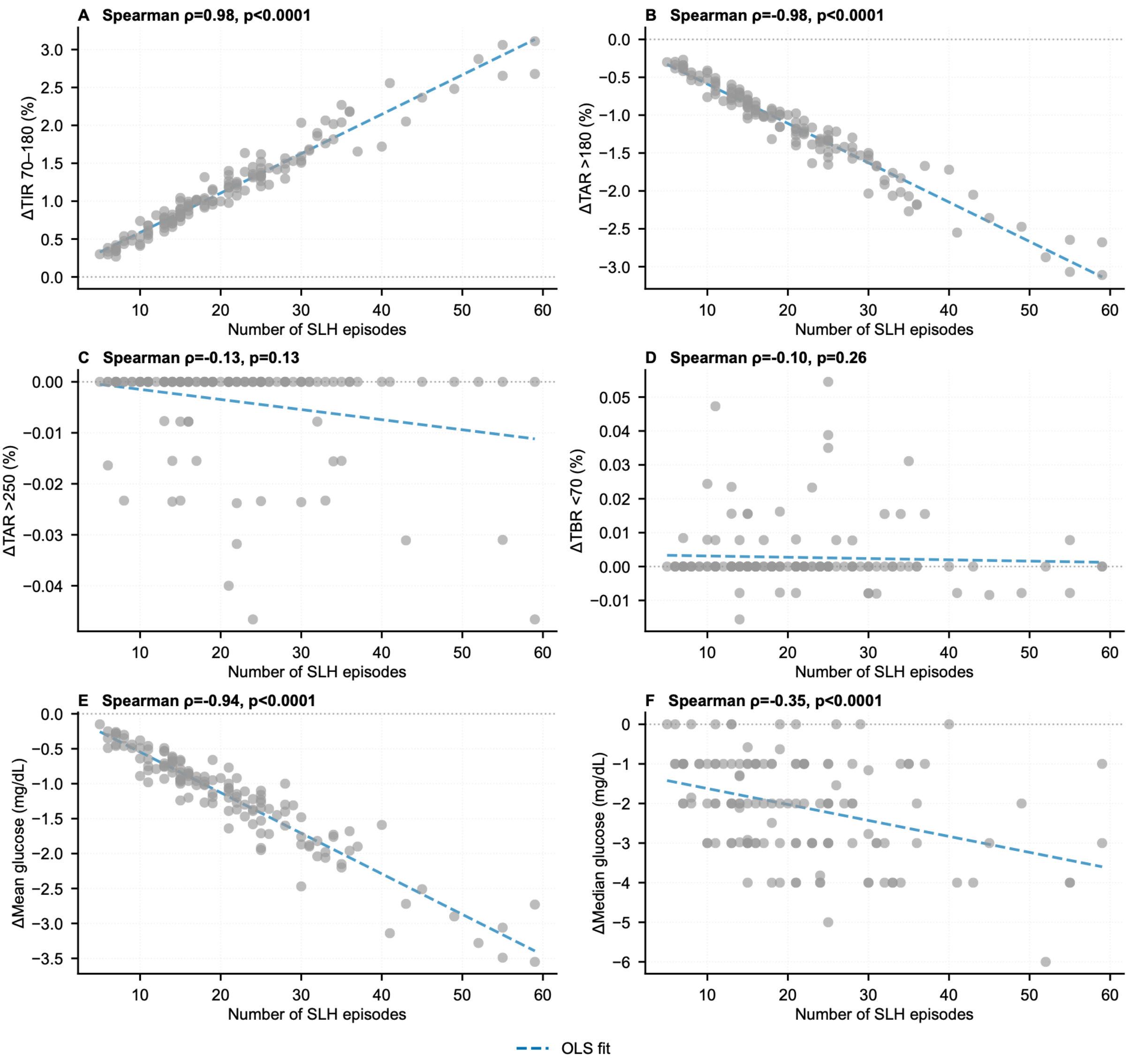


**Figure S9:** Change in each CGM summary metric after in silico removal of short-lived hyperglycaemia (SLH) episodes, against the number of SLH episodes. Each point is one participant (n=135): (A) ΔTIR 3·9–10·0 mmol/L, ρ = 0·98, p<0·0001; (B) ΔTAR >10·0 mmol/L, ρ = −0·98, p<0·0001; (C) ΔTAR >13·9 mmol/L, ρ = −0·13, p=0·13; (D) ΔTBR <3·9 mmol/L, ρ = −0·10, p=0·26; (E) ΔMean glucose, ρ = −0·94, p<0·0001; (F) ΔMedian glucose, ρ = −0·35, p<0·0001 (percentage points for TIR, TAR and TBR; mmol/L otherwise; ρ: Spearman correlation). Dashed line: ordinary least-squares fit; dotted line: no change. Counterfactual signals (δ=30 min) were built as described in Supplementary methods S1; metrics were computed over the whole day.

### Figure S10

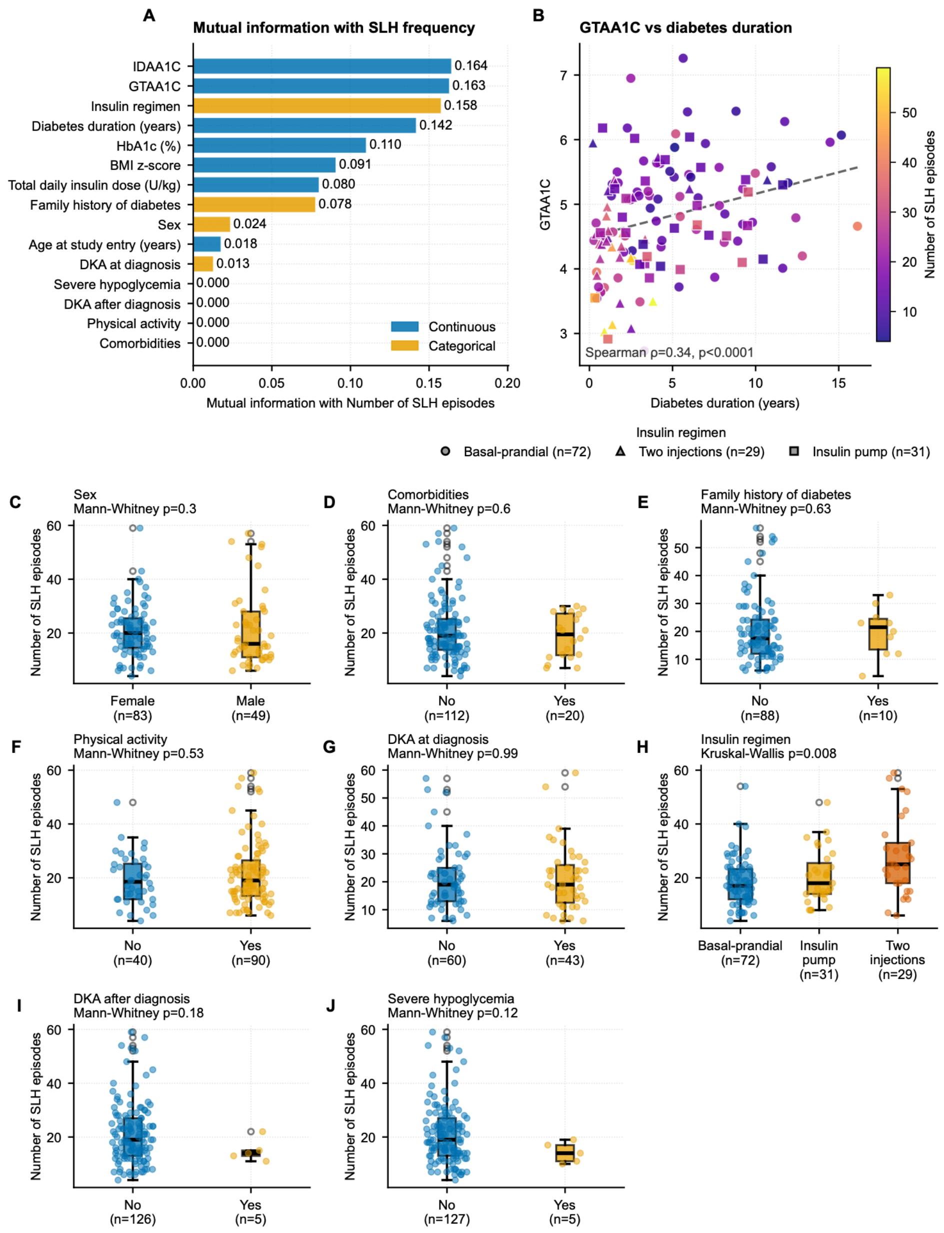


**Figure S10:** Patient-phenotype determinants of short-lived hyperglycaemia (SLH) frequency.(A) Mutual information between each phenotype variable and the number of SLH episodes per participant (categorical variables one-hot encoded, per-category values summed); the three highest were IDAA1c, GTAA1c, and Insulin regimen. (B) GTAA1c against diabetes duration, coloured by SLH count and shaped by insulin regimen; dashed line: least-squares fit (Spearman ρ = 0·34, p<0·0001). (C–J) Number of SLH episodes by category for each categorical phenotype variable. Boxes: median and IQR; whiskers: 1·5 × IQR; dots: individual participants. Group sizes and p values (Mann–Whitney for two groups, Kruskal–Wallis for three or more; uncorrected) are shown in each panel.
